# Exploratory comparison of Healthcare costs and benefits of the UK’s Covid-19 response with four European countries

**DOI:** 10.1101/2020.12.14.20248201

**Authors:** Howard Thom, Josephine Walker, Peter Vickerman, Will Hollingworth

## Abstract

**Background:** In responding to covid-19, governments have tried to balance protecting health while minimising Gross Domestic Product (GDP) losses. We compare health-related net benefit (HRNB) and GDP losses associated with government responses of the UK, Ireland, Germany, Spain, and Sweden from UK healthcare payer perspective.

**Methods:** We compared observed cases, hospitalisations, and deaths under “mitigation” to modelled events under “no mitigation” to 20^th^ July 2020. We thus calculated healthcare costs, quality adjusted life years (QALYs), and HRNB at £20,000/QALY saved by each country. On per population (i.e. per capita) basis, we compared HRNB with forecast reductions in 2020 GDP growth (overall or compared to Sweden as minimal mitigation country) and qualitatively and quantitatively described government responses.

**Results:** The UK saved 3.17 (0.32-3.65) million QALYs, £33 (8-38) billion healthcare costs, and £1416 (220-1637) HRNB per capita at £20,000/QALY. Per capita, this is comparable to £1,455 GDP loss using Sweden as comparator and offsets 46.1 (7.1-53.2)% of total £3075 GDP loss.

Germany, Spain, and Sweden had greater HRNB per capita. These also offset a greater percentage of total GDP losses per capita. Ireland fared worst on both measures. Countries with more mask wearing, testing, and population susceptibility had better outcomes. Highest stringency responses did not appear to have best outcomes.

**Conclusions:** Our exploratory analysis indicates the benefit of government covid-19 responses may outweigh their economic costs. The extent that HRNB offset economic losses appears to relate to population characteristics, testing levels, and mask wearing, rather than response stringency.

**Research in Context:** *Evidence before this study:* Our research question was how the health-related net benefits and economic impacts of the UK response to the covid-19 epidemic first wave compared to other European countries. We searched PubMed, MedRxiv, and Arxiv for terms related to cost-effectivness, covid-19, and the UK. Two studies compared predicted lives saved to predicted gross domestic product (GDP) losses. One found that lives saved by a lockdown would outweigh GDP losses, while another found a lockdown to cost £10million per life saved. A later modelling study used quality adjusted life-years (QALYs), going beyond lives saved, and found cost per QALY was below £50,000. A fourth, comparing observed to modelled deaths and hospitalisations, found the cost per QALY was at least £220,000, and thus the UK response was not cost-effective. None of these were international comparisons. One international study found good health and economic outcomes to be correlated. Another found global trade reductions and voluntary behavioural changes to have greater impact on economic growth than government measures. Neither considered cost-effectiveness. However, they suggest comparison to GDP loss is naïve as this is total loss and not that due to government restrictions.

*Added value of this study:* We compare the UK to Ireland, Germany, Spain, and Sweden on health-related net benefits and economic impacts of government response from a UK National Health Service (NHS) perspective. We describe countries’ response measures. We compared model predictions of outcomes under “no mitigation” to observed outcomes under “mitigation” up to July 20^th^ 2020. We estimated healthcare costs, QALYs, and health-related net benefit (HRNB) saved. We compared HRBN to GDP losses using Sweden as a “minimal mitigation” comparator and calculated the % of total GDP loss they offset. We found the UK saved 3·17 (0·32-3·65) million QALYs, £33 (£8-38) billion in healthcare costs, and £1416 (220-1637) HRNB per capita at the NHS threshold of £20,000/QALY. This is comparable to the £1,455 GDP loss per capita using Sweden as comparator and offsets 46·1% (7·1-53·2) of the total estimated £3075 GDP loss per capita. We found that Germany, Spain, and probably Sweden had greater HRNB per capita and offset greater percentages of GDP loss per capita. Ireland fared worst on both measures. We found countries with more mask wearing, testing, and population susceptibility (e.g. older and more interpersonal contact) had better outcomes. Highest stringency responses did not appear to have best outcomes.

*Implications of all the available evidence:* We add to growing evidence that the total economic impact of covid-19 exceeds the HRNB of the UK’s response. However, using Sweden as comparator and comparing across countries, we argue that GDP loss is not purely due to government restrictions and that due to restrictions may be outweighed by HRNB. We evaluated the extent to which countries have offset GDP losses, and these appear to be higher in countries with more at-risk populations, higher testing, and higher mask wearing, rather than those with most stringent restrictions.

## Introduction

Covid-19 has caused severe health and economic damage since its emergence in Wuhan, China at the end of 2019. As of 13th November 2020, the World Health Organisation (WHO) has reported over 50 million confirmed cases and 1.29 million deaths globally.^1^ In the UK there have been over a million cases and over 60,000 deaths. The economic damage has been similarly dramatic with the International Monetary Fund (IMF) reducing its forecast Gross Domestic Product (GDP) growth for 2020 in the UK from 1.4% in January to −9.8% in October.^2,3^ Similar reductions have been observed across Europe and the world. Health system responses (e.g. test and trace), government recommended or mandated social distancing measures, and financial assistance have so far been the primary response.^4^

The economic case for stringent social distancing measures has been questioned due to their impact on the economy and livelihoods.^5,6^ A cost-benefit analysis of government responses to covid-19 requires an infectious disease model to project the counterfactual of what would have happened in the absence of government intervention. The first wave of covid-19 in Europe was largely complete by July 2020; complete data on observed outcomes are therefore available for comparison with modelled outcomes, and it is thus possible to compare the costs and benefits of Government responses to this first wave.^1^ The costs of social distancing measures are largely borne by society and the economy. A potential approach is to simply compare health-related net benefits (health gains minus healthcare costs) to losses in GDP. However, this would underestimate the value of government intervention on social distancing as some GDP losses are due to covid-19 ill-health, reductions in global trade, and voluntary changes in behaviour, rather than mandatory social distancing measures. Some studies have found that the much of the reduction is due to global and voluntary changes.^7,8^

Zala 2020 was an early UK modelling study aiming to assist policy makers but not making use of observed data. They found that even strategies causing a 10% reduction in national income are cost-effective at £50,000 per QALY.^9^ Lifetables and age-specific QALY norms were used to estimate that 8.8 QALYs were lost for each covid-19 death, similar to estimates by Briggs 2020.^10^ Miles 2020 compared predictions of outcomes under no mitigation to observed outcomes up to June 2020, using an informal estimate of 5 QALYs lost per death.^6^ They compared the reductions in actual GDP growth forecasts to costs and QALYs saved by government response. They found that continuing the lockdown was only cost-effective at a willingness-to-pay per QALY between £220,000 and £3.7 million.

Building on this earlier work, we aim to quantify the health-related net benefit of Government interventions and compare them to the loss in GDP associated with social distancing measures in five western European countries over the first wave of covid-19. We use a new model of cost and QALYs of covid-19 cases, hospitalisations, and deaths, with an estimate of QALYs lost per death. We compare observed outcomes to new forecasts under no mitigation using an adaptation of a published open-source covid-19 model.^11^ Multiple forecasts are considered to explore uncertainty. Health-related net benefits are compared per capita with published reductions in 2020 GDP growth forecasts, both in total and using Sweden as a minimal mitigation comparator. Differences are presented alongside a description of social distancing and other measures, allowing us to illustrate characteristics of a successful response.

## Methods

The comparisons cover the period 1^st^ January to 20^th^ July 2020, corresponding to the first wave of covid-19.^1^ The “mitigation” strategy is based on observed disease outcomes while “no mitigation” is based on modelled projections.

### Comparator countries and description of response

Comparator countries were selected as they were high income Western European healthcare systems with a range of responses and outcomes to covid-19. Sweden had a less stringent response than the UK, Germany had lower case and death rates, Ireland took earlier and more stringent action, and Spain had similar case and death rates.^1,12^ We reviewed each country’s response with reference to Wikipedia, news reports, and government websites.^13^ Government responses were categorised under international travel, mass gatherings, pubs/restaurants, education, stay-at-home measures, financial assistance, mask requirements, and testing. Timings were relative to the first confirmed covid-19 case and death in each country. As numbers of cases and tests are likely correlated, we report tests per covid-19 death. For additional comparison we used the maximum and average, up to 20^th^ July, University of Oxford Stringency Index for each country.^12^ This composite measure is based on nine response indicators, including school closures, workplace closures, and travel bans.

### Observed outcomes under “mitigation”

Government and news websites from each of the countries were searched for data on the numbers of covid-19 cases, hospitalisation, and deaths that took place from 1^st^ January to 20^th^ July 2020.Covid-19 related deaths rather than excess deaths were used as these were available for all countries of interest.

### Modelling outcomes under “no mitigation”

We conducted a search for existing models on PUBMED, the Arxiv and medRxiv preprint servers. We selected the London School of Hygiene and Tropical Medicine Centre for the Mathematical Modelling of Infectious Diseases (CMMID) covid-19 model (version 1) for projections in all countries of interest. This is an open-source age-structured deterministic mathematical model of SARS-CoV-2 transmission.^11,14,15^ This model was chosen because it included projections for all countries of interest, allowed common structural assumptions between countries, and enabled tailoring of virus introduction dates and reproduction numbers (*R*_0_). It accounts for differences in susceptibility and symptomatic rate by age, with children less susceptible to infection and more likely to be asymptomatic than adults.

We used results of meta-analysis studies and preprints published before Feb 26, 2020 to estimate the “no mitigation” *R*_0_ at 2.7 (95% credible interval 1.6-3.9).^14^ Our scenario A uses 2.7 while scenarios B and C assume *R*_0_ to be 1.6 and 3.9, respectively. This *R*_0_ point estimate was sufficiently early to avoid the influence of public health interventions. Other modelling studies have used similar estimates of 2.6 (uncertainty range: 1.5-3.5) and 3.8 and there is limited evidence of variation between western European countries. ^16–18^ Default values were used for other parameters. The model forecasts for each *R*_0_ scenario in each country were then downloaded including cumulative cases, ICU bed days, non-ICU bed days, and deaths from virus introduction to 20^th^ July 2020. Bed days were converted to numbers of admissions using the duration of non-ICU and ICU stays assumed by CMMID. These values and further modelling details are in the Appendix.

### Calculating health-related net benefit of mitigation

Our calculations include QALYs lost from covid-19 cases, hospitalisations, and deaths. We include only direct health impacts, and not indirect impacts such as mental health effects of bereavement or social distancing. We used influenza quality of life decrements for hospitalised or non-hospitalised cases who survive as estimates are not yet available for covid-19 (values in Appendix). We estimate QALYs lost per covid-19 death using country specific distributions of age at death, life expectancy, and age-specific quality of life norms (details in Appendix).^10^ Our baseline estimate of QALYs lost assumes an SMR of 1.1 (see Appendix) but there is uncertainty about the prevalence of comorbidities in those dying from covid-19 compared to the general population. ^19^ We therefore consider a sensitivity analysis with a high SMR of 2.0.

### Health-related costs of treatment

We estimated hospitalisation costs in 2020 UK £ pounds sterling from a UK NHS perspective. We assumed no healthcare cost for community covid-19 cases as prescription medications typically have low cost and over the counter medications are not funded by the NHS. Using the figures for hospital stay costs, percentage requiring high dependency or ICU, average days ICU, and cost per day of ICU gives an average cost of £4847 per hospitalized patient (details in Appendix). We did not include costs for testing or tracing as the level under “no mitigation” is unknown and under “mitigation” was not reported by all countries.

### Calculating direct health-related net benefit of mitigation

The total QALYs lost and healthcare costs were estimated for the observed “mitigation” and modelled “no mitigation” strategies. Incremental QALYs gained and healthcare savings from mitigation were calculated. A monetary incremental health-related net benefit was calculated by multiplying incremental QALYs by the conventional UK willingness-to-pay threshold of £20,000/QALY and subtracting incremental costs.^20^ These are presented per capita using 2020 population estimates.^21^

### Economic impact

We used the October 2020 International Monetary Fund (IMF) World Economic Outlook projections for GDP growth in 2020 to give estimates of growth with the covid-19 pandemic and compared these to the corresponding January 2020 pre-pandemic projections.^3,22^ January IMF estimates for Sweden and Ireland were not available so instead Organisation for Economic Cooperation and Development (OECD) 2020 projections from 21^st^ November 2019 were used.^23^ We calculated £ pound sterling value of GDP changes using IMF estimates of total GDP value in October 2019 and OECD 2019 purchasing power parities. ^2,24^ Final results are reported per capita using 2020 population estimates.^21^

Only part of the observed GDP loss is a result of government mandated social distancing measures, with the rest a consequence of global trade changes, voluntary behaviour changes, and productivity loss due to ill health. We therefore present two scenarios. The first assumes that under no mitigation the 2020 GDP reduction would have been the same as that observed in Sweden. Sweden might be considered to represent the “minimal mitigation” policy that is politically acceptable in a western European democracy during a global pandemic, rather than “no mitigation”. In the second we estimate the percentage of observed GDP loss that was offset by net QALYs gained and healthcare savings.

## Results

### Description of country responses to covid-19

Apart from Sweden, response measures and timings were similar across countries (full details in Appendix). Ireland banned gatherings 15-28 days earlier than other countries. All countries except Sweden ordered pubs and restaurants to shut but Ireland did so up to 40 days earlier. Spain, Germany and the UK closed schools at a similar timepoint, roughly 30 days after Ireland. Sweden recommended secondary schools and universities to move to distance learning but kept nurseries and schools open. The UK introduced stay-at-home orders almost 30 days later than Ireland and 12 days after Spain. Sweden had no stay-at-home order. All countries introduced wage support at similar timepoints. Requirements for mask wearing were eventually introduced for all countries except Sweden, but Spain and Germany moved much earlier than the UK and Ireland. Most countries did not introduce comprehensive tracing until the very end of the first wave. Testing levels per death were much higher in Germany, with 812.8 compared to 324.6 for Ireland with the second highest level. According to the stringency Index, the UK had lower maximum level restrictions than Ireland and Spain but higher than Germany and Sweden. Ireland had the highest maximum level of restrictions. Average restriction index was very similar between countries other than Sweden.Sweden had the least stringent measures (maximum 64.81 vs next lowest 76.85 and average 38.79 vs next lowest 45.41).

### Comparison of responses to covid-19 on cases, hospitalisations, and deaths

Differences in prevented covid-19 cases, hospitalisations, and deaths under “mitigation” are illustrated in Figure 1, with full details in the Appendix.

**Figure 1.**
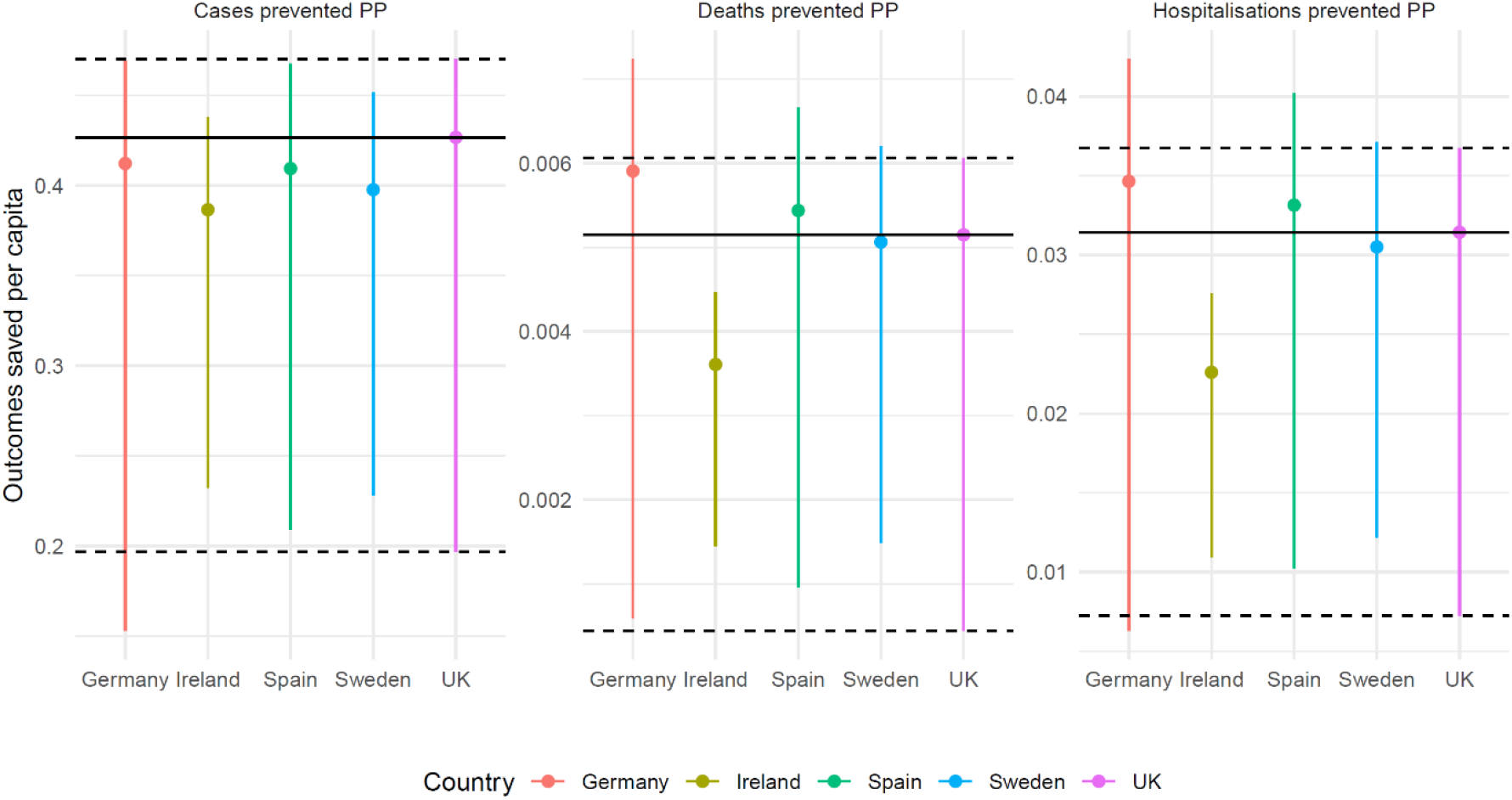
Outcomes saved per capita (PP). Central estimate is scenario A while lower and upper limits correspond to scenarios B and C, respectively. Horizontal solid lines indicate UK estimates for ease of comparison. Values above these lines indicate greater benefit per capita. Comparison of cases is affected by levels of testing so focus should be on deaths and hospitalisations.

Observed case numbers are affected by variance in levels of testing across countries limiting comparability. Hospitalisations and deaths per capita were highest in Sweden, Spain, and the UK. Ireland and Germany had much lower hospitalisations and deaths.

Under base case scenario A (*R*_0_=2.7), Spain and Germany prevented more hospitalisations and deaths per capita than the UK, while Sweden was similar to the UK. Ireland prevented fewer hospitalisations and deaths than all countries, likely driven by a younger population. Patterns are similar under scenario C (*R*_0_=3.9) but reverse for Germany and Ireland under A (*R*_0_= 1.6); this nonintuitive reversal is due to a nonlinear relationship between *R*_0_ and outcomes.

### Comparison of health-related costs, benefits, and net benefits

The incremental health-related benefits (QALYs), costs, and per capita incremental net benefits are presented in Table 1. In scenario A (*R*_0_=2.7), health-related net benefits per capita are highest for Germany, followed by Spain, UK, and Sweden. Ireland had almost 30% lower net benefit than Sweden. These findings are driven by prevented hospitalisations and deaths (Figure 1).

**Table 1.** Health-related benefits, costs, and net benefits of the government response for SMR=1.1 scenarios. Scenarios are for the “No mitigation” simulations from the CMMID model which are compared to the observed outcomes. Comparisons up to 20^th^ July 2020. (INB = Incremental health-related net benefit).

| Scenario | Country | Incremental benefit<br>(millions of QALYs) | Incremental costs<br>(£Billions) | INB per capita at<br>£20,000/QALY (£) |
| --- | --- | --- | --- | --- |
| A<br>$R_0 = 2.7$ | UK | 3.166 | -32.94 | 1,416.6 |
|  | Ireland | 0.162 | -1.67 | 994.6 |
|  | Spain | 2.297 | -23.84 | 1,492.6 |
|  | Germany | 4.467 | -43.22 | 1,581.2 |
|  | Sweden | 0.463 | -4.78 | 1,388.2 |
| B<br>All $R_0 = 1.6$ | UK | 0.319 | -8.61 | 220.5 |
|  | Ireland | 0.071 | -0.82 | 453.3 |
|  | Spain | 0.452 | -8.00 | 364.6 |
|  | Germany | 0.501 | -8.03 | 215.3 |
|  | Sweden | 0.149 | -2.04 | 495.4 |
| C<br>All $R_0 = 3.9$ | UK | 3.649 | -38.28 | 1,637.4 |
|  | Ireland | 0.197 | -2.04 | 1,206.6 |
|  | Spain | 2.754 | -28.75 | 1,792.6 |
|  | Germany | 5.357 | -52.79 | 1,907.6 |
|  | Sweden | 0.555 | -5.77 | 1,668.1 |

Under scenario B (*R*_0_=1.6), Sweden and Ireland have the greatest health-related benefit while the UK and Germany have the lowest, again driven by nonlinear modelling relationship between *R*_0_ and outcomes (Figure 1). Under scenario C (*R*_0_=3.9), almost the same pattern as the base case is found. Patterns are the same under SMR=2.0, but health-related net benefits are marginally lower (see Appendix). Deaths account for 60-70% of incremental health-related net benefits, with the remainder due to hospitalisations (Appendix). Over 95% of the benefit of prevented hospitalisations is due to costs, rather than QALYs.

Across all scenarios, with *R*_0_=2.7 as base case, the UK response is estimated to have saved 3.17 million (ranging from 0.32 to 3.65 million) QALYs, £33 billion (£8-38 billion) in healthcare costs, and gained £1416 (220-1637)per capita health-related net benefit at £20,000/QALY.

### Comparison of economic impact of responses to covid-19

Estimates of the economic impact of government responses are presented in Table 2. Spain and the UK had the worst GDP loss, Germany and Ireland had much smaller reductions in GDP. Despite high hospitalisation and deaths, Sweden had the lowest reduction in GDP, possibly explained by lower severity restrictions.

**Table 2.**
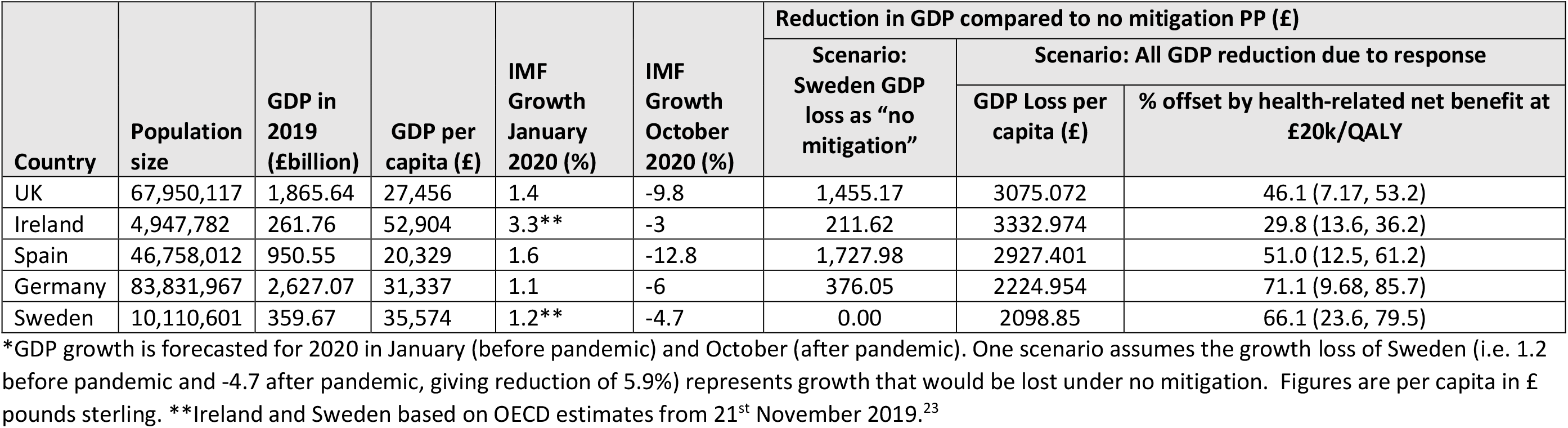
International comparison of GDP growth forecasts, losses due to the pandemic and government response, and % offset by health-related net benefit.

| Country | Population size | GDP in 2019 (£billion) | GDP per capita (£) | IMF Growth January 2020 (%) | IMF Growth October 2020 (%) | Reduction in GDP compared to no mitigation PP (£) |  |  |
| --- | --- | --- | --- | --- | --- | --- | --- | --- |
|  |  |  |  |  |  | Scenario: Sweden GDP loss as “no mitigation” | Scenario: All GDP reduction due to response |  |
|  |  |  |  |  |  |  | GDP Loss per capita (£) | % offset by health-related net benefit at £20k/QALY |
| UK | 67,950,117 | 1,865.64 | 27,456 | 1.4 | -9.8 | 1,455.17 | 3075.072 | 46.1 (7.17, 53.2) |
| Ireland | 4,947,782 | 261.76 | 52,904 | 3.3** | -3 | 211.62 | 3332.974 | 29.8 (13.6, 36.2) |
| Spain | 46,758,012 | 950.55 | 20,329 | 1.6 | -12.8 | 1,727.98 | 2927.401 | 51.0 (12.5, 61.2) |
| Germany | 83,831,967 | 2,627.07 | 31,337 | 1.1 | -6 | 376.05 | 2224.954 | 71.1 (9.68, 85.7) |
| Sweden | 10,110,601 | 359.67 | 35,574 | 1.2** | -4.7 | 0.00 | 2098.85 | 66.1 (23.6, 79.5) |

\*GDP growth is forecasted for 2020 in January (before pandemic) and October (after pandemic). One scenario assumes the growth loss of Sweden (i.e. 1.2 before pandemic and -4.7 after pandemic, giving reduction of 5.9%) represents growth that would be lost under no mitigation. Figures are per capita in £ pounds sterling. \*\*Ireland and Sweden based on OECD estimates from 21<sup>st</sup> November 2019.<sup>23</sup>

In scenario A, observed GDP loss exceeded health-related net benefits in all countries (Table 1 and Table 2). The extent to which GDP losses were ‘offset’ by net health benefits ranged widely from a low of 30% in Ireland to 71% in Germany at the £20,000/QALY threshold (Table 2). These findings were very sensitive to assumptions about *R*_0_. For example, in the UK the extent to which GDP loss is offset was 46% in scenario A but ranged from 7% to 53% (Table 2). If, however, the 5.9% GDP loss observed in Sweden is considered the minimal level for European economies, given the effects of global trade and voluntary behavioural changes, then under scenario A the net health benefits in Germany and Ireland exceed the GDP loss whereas they fall short but are comparable in the UK and Spain. The analysis with SMR=2.0 (Appendix) reduces net benefits per capita but does not change our overall findings.

Figure 2 illustrates that in countries (Ireland, Spain, and the UK) with higher social distancing stringency, net health benefit did not offset a greater proportion of GDP loss. It illustrates that in Germany, with highest testing per death, net health benefits offset the greatest % GDP loss. It illustrates that in Ireland, with lowest predicted “no mitigation” death rate, net health benefits of mitigation offset the lowest % GDP loss. It also suggests that countries with lower median age (e.g. Ireland) had lower offsets than those with higher age (e.g. Germany). None of these patterns are consistent and there is insufficient data for statistical testing.

**Figure 2.**
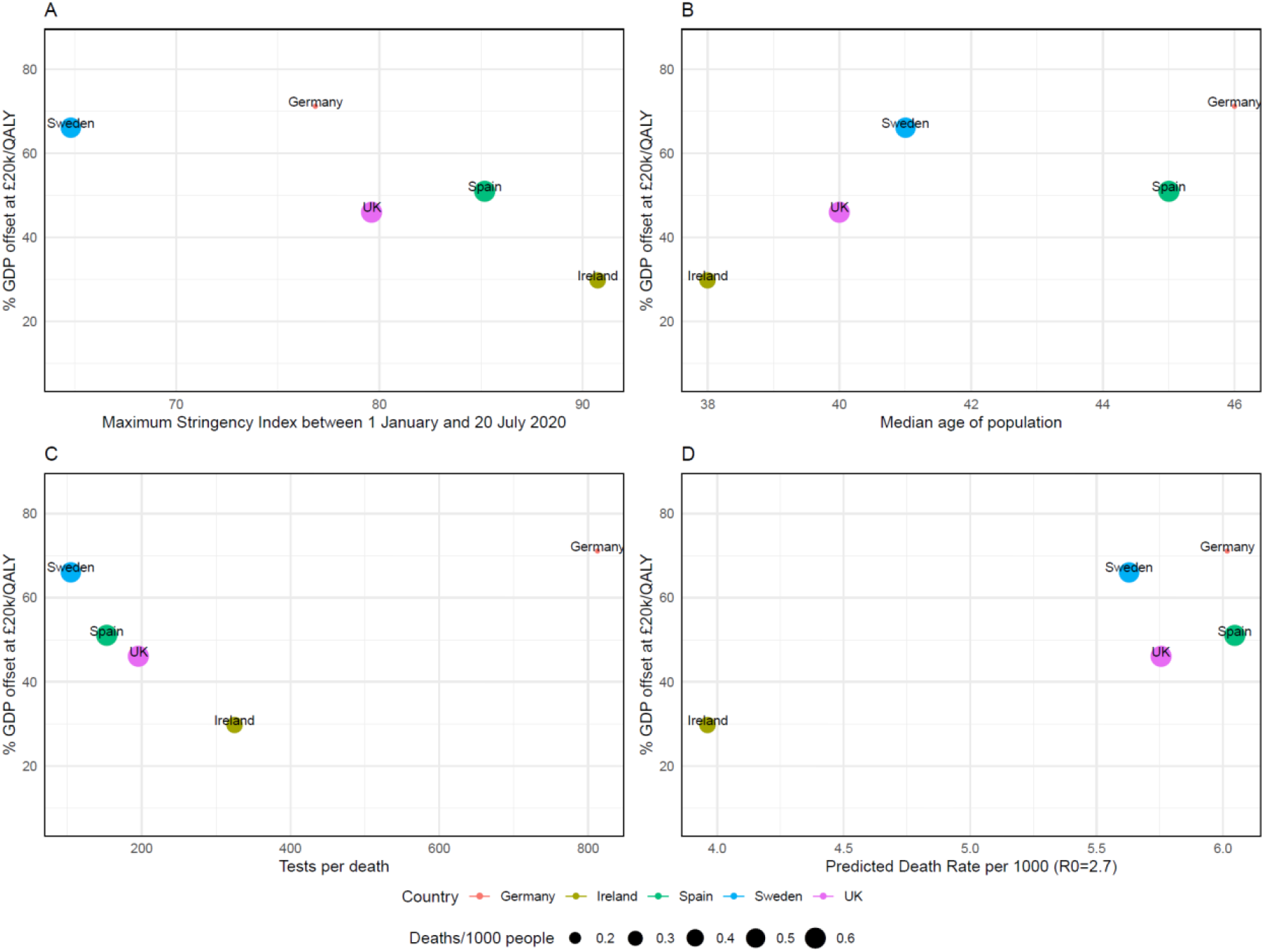
Comparison of % GDP loss offset by net health benefits at £20,000/QALY across countries and comparison with explanatory factors. A) maximum stringency index, B) median age of population^21^, C) tests per death, D) predicted death rate under no mitigation scenario.

### Discussion

We have provided an exploratory comparison of health-related benefits and costs saved by government measures across European countries. Mitigation saved lives and reduced healthcare treatment costs substantially, with the health-related net benefit in the UK being £1416 (220-1637) per capita. In all countries, projected GDP loss is higher than health-related net benefits but this ignores that GDP would have reduced in the absence of government mandated social distancing measures. In the UK, 46.1 (7.1-53.2)% of GDP loss was offset by health-related net benefit, but higher offset percentages were estimated in Germany (71.1%), Spain (51.0%), and Sweden (66.1%). Countries with tighter social distancing restrictions do not appear to perform better on this measure. Instead, the distinguishing features of Germany (best performing country) are higher testing, earlier mask wearing, and higher median age. Germany having the highest % GDP offset may also be explained by having lowest absolute death and hospitalisation rates.^7^ The defining feature of Ireland (worst performing country) is low likelihood of covid-19 hospitalisation and death, possibly explained by lowest median age (38 years).^25^ High % offset countries Germany and Spain may simply have had older populations with more to benefit (Figure 2). Sweden had lower levels of testing, no requirement for mask wearing, and mostly voluntary but had similar health-related benefits to the UK; median age (41 years) is similar to the UK (40 years) again suggesting population susceptibility is a key determinant of success.

There are significant limitations to our analysis. There is limited understanding of plausible parameter ranges (e.g. *R*_0_) and limitations of using an existing model so we could not conduct probabilistic sensitivity analysis. Costs of hospitalisation were assumed to be independent of the number of hospitalisations; however, once capacity is breached, additional facilities would need to be constructed at substantial cost. We did not model testing or tracing costs. The UK has allocated £22 billion, or £338 per capita, on testing and tracing over 2020-21 so this could become important for future evaluations.^26^ We did not model indirect health impacts, such as the impact of either social distancing or a more severe pandemic on mental health or delivery of healthcare. There are limitations in the extent to which any single measure of stringency can capture complex Government policies. For example, Finland had a maximum of 67.6 and average of 36.6 compare to 64.8 and 38.8 in Sweden, respectively, despite introducing a strict lockdown.^12^ The CMMID model represents a worst case scenario for “no mitigation” as it does not include voluntary changes in behaviour. Finally, our restriction to the first wave of the pandemic means we may be comparing cases, hospitalisations, or deaths postponed, rather than prevented, and we take no account for possible herd immunity.

Estimating only the impact of government social distancing measures on GDP is difficult. Baker 2020 estimated that at least half of economic impact is due to global uncertainty, while Chen-2020 found that outbreaks and voluntary mobility reductions have a much greater impact.^8,27^ Sweden represents “minimal mitigation” rather than “no mitigation”. Furthermore, Sweden’s neighbouring countries introduced greater restrictions and also achieved lower forecast reductions in GDP (e.g. Norway with maximum stringency index 79.6 and forecast −4.0% growth, Denmark with maximum index 72.2 and −4.5% growth).^12,22^ It is clear that there is no simple trade-off between economic pain and health gains. Finally, comparisons for Ireland may be skewed as its GDP per capita is subject to disproportionate globalisation effects.^28^

A challenge to our % GDP loss offset is that the £20,000/QALY threshold relates to NHS expenditure, not society. Societal thresholds may range from £10,000/QALY to £70,000/QALY.^9^ The NHS uses a threshold of £50,000/QALY for patients at end-of-life, possibly relevant to patients experiencing severe covid-19.^20^ Furthermore, the UK uses higher thresholds when there is perceived public pressure, as in the case of the Cancer Drugs Fund with an estimated threshold of £220,000/QALY.^29^ The UK’s benefits would outweigh the total GDP loss at above £70,000/QALY (Appendix).

There are possible lessons from our analysis for the ongoing covid-19 pandemic. One is to consider the cost and QALYs associated with hospitalisations and deaths, rather than only total numbers, when making decisions. Successful responses appear to be linked to higher levels of testing or mask wearing, rather than strictness of response. The experience of Ireland suggests that consideration should then be given to the maximum potential benefit, which depends on demographics and interpersonal contacts. This final consideration could be applied regionally rather than nationally, to better target approaches to future outbreaks.

## Conclusion

Our exploratory analysis estimates that the UK saved 3.17 million (0.32 to 3.65 million) QALYs, £33 billion (£8-38 billion) in healthcare costs, and £1416 (220-1637) health-related net benefit at £20,000/QALY per capita. This is comparable to £1,455 GDP loss per capita using Sweden as “minimal mitigation”, while it is 46.1 (7.1-53.2)% of the total £3075 GDP loss per capita. Germany, Spain, and, in most scenarios, Sweden had greater health-related net benefits per capita and less GDP loss per capita. Ireland fared worst on both measures.

At face value, the total economic impact of covid-19 exceeded the health-related net benefits. However, it is not realistic to attribute the full economic impact solely to government responses or to argue that any European country could have avoided their GDP loss or applied “no mitigation”. We have attempted to evaluate the extent to which economic costs have been offset by net health benefits. Countries with susceptible populations, higher testing, and higher mask wearing, rather than those with the most stringent restrictions appear to have done better on this measure.

## Data Availability

Our model is implemented in the R statistical programming language. Parameters and data are in a Microsoft Excel spreadsheet. Code and data are publicly available: https://github.com/Bogdasayen/covid_cea

## Funding and acknowledgments

We thank Sam Abbott at LSHTM for his advice on the our modelling assumptions. Funding was provided by the Elizabeth Blackwell Institute Rapid Response Call (COVID-19) 2020, Medical Research Council grant MR/S036709/1, and National Institute for Health Research Bristol Biomedical Research Centre at University Hospitals Bristol and Weston NHS Foundation Trust and the University of Bristol.

## Conflicts of interest

Authors have no conflicts of interest to declare.

### Key points

1. We estimate the UK government response to covid-19 up to 20^th^ July saved 3.17 (0.32-3.65) million quality adjusted life years (QALYs), £33 (8-38) billion healthcare costs, and £1416 (220-1637) health-related net benefit (HRNB) per capita at £20,000/QALY.
2. Per capita, this HRNB is comparable to a £1,455 gross domestic product (GDP) loss using Sweden as comparator and offsets 46.1 (7.1-53.2)% of total £3075 GDP loss.
3. Germany, Spain, and Sweden had greater HRNB per capita and offset a greater percentage of total GDP losses per capita than the UK, while Ireland fared worse on both measures.
4. Countries with susceptible populations, higher testing, and higher mask wearing, rather than those with the most stringent restrictions, appear to have had greatest HRNB and offset the greatest percentage of total GDP losses.

## Appendix A. Modelling details

### A.1 Search details

We used the preprint server Arxiv application programming interface (API) to search for hits with the terms “COVID”, “CoV”, or “coronavirus” in their titles or abstracts. We then filtered these 1432 hits (19-June-2020) by those containing “model” and either “United Kingdom”, “Great Britain”, or “England” in their abstracts. This gave 5 for abstract/full-text screening. We applied the same search strategy to medical preprint server medRxiv, which gave 5387 hits (18-June-2020), and 61 for title/abstract screening, of which only 9 were relevant for full-text screening. We also conducted a PUBMED search for titles or abstracts containing terms related to covid-19 (“coronavirus”, “SARS-CoV-2”, “covid-19”), the UK (“United Kingdom”, “England”,”UK”, “Great Britain”), and the string “model”. This identified 64 hits, of which 7 were relevant for full-text screening. We supplemented these targeted literature searches with reviews of websites for established modelling groups in the UK, namely the Medical Research Council (MRC) Centre for Global Infectious Disease Analysis

Imperial College London, the Nuffield Department of Health at University of Oxford, and the Centre for the Mathematical Modelling of Infectious Diseases (CMMID) at the London School of Hygiene and Tropical Medicine. We also searched the Society for Medical Decision Making (SMDM) database of available models (as of 18-June-2020); this included 14 potentially relevant models but all were non-UK except those already identified in the medRxiv searches. We used the same search strategy to identify models of transmission in each of the countries of interest.

### A.2 Further details on use of CMMID model for projections under “no mitigation”

In all modelling scenarios, we calibrated the virus introduction date such that the modelled first death date (assumed to be the first date when total modelled deaths were greater than 0.5, as the model estimates fractional deaths) matched to observed data on first death in each country (details in Appendix). We also kept the default contact matrices for home, work, school, and ‘other’ settings; UK matrices were based on a 2008 study which used questionnaires and contact diaries to measure contact patterns in the UK and 7 other European countries (POLYMOD), while non-UK matrices were based on 2013 and 2017 studies which extrapolated from the results of the POLYMOD study.^1,2^

Other than our changes to the *R*_0_ value, we used the default parameter values. These included risk of death and hospitalization not modified from baseline age-dependent parameters, and 10 initial infections on virus introduction date) with the baseline (no mitigation) scenario being used for our analysis.

### A.3 Further modelling methods – estimating QALYs lost per death

At the time of submission, the paper of Briggs was not fully published.^3^ For completeness, we therefore reproduce the key steps used from Briggs’ analysis to estimate QALYs lost per death.

Assuming an initial cohort of 100,000 births, the number surviving to age x years is given by

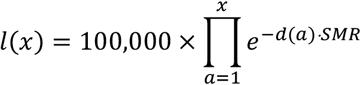

Where *d*(*x*) = −*ln*{1 − *q*(*x*)} is the hazard rate of death corresponding to the probability of death q(x) for period *x* to *x* + 1, and *SMR* is a standardized mortality ratio for our cohort (i.e. people dying of covid-19). The *SMR* for patients dying from covid-19 is unknown. Public Health England report that the percentage of covid-19 deaths with comorbidities were similar to deaths in the general population. ^4^ The greatest disparities were in diabetes (6.5% difference: 14.6% vs 21.1%) and hypertensive diseases (4.1% difference: 14.5% vs 19.6%).^4^ We combine these two to give ~10% of patients with comorbidities and take a high bound of 2.0 for the conditions’ standardised mortality ratios (SMR), giving a population average SMR of 1.1. We also consider an extreme sensitivity with SMR of 2.0.

We average the *l*(*x*) over males and females assuming simplistically that the proportion of females is 0.5. The number of person-years lived between ages *x* and *x* + 1 for x ≥ 1 is

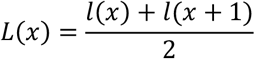

This is the duration of the interval (1 year in this case) times the average number alive during the interval. The life expectancy of people surviving to age x is then the expected number of life years of cohort aged *x* divided by number surviving to age *x* (i.e. average number of years of life)

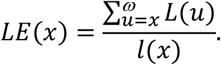

The *ω* is some upper limit on life expectancy, which we take to be 100 years. Life years are then weighted by their age and country specific QALY norms *Q*(*u*) for age *u* to give quality adjusted life expectancy (QALE).

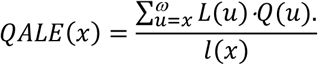

The final step is to discount at rate *r* giving the discounted QALYs lost per death at age x

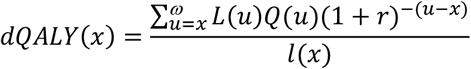

This is calculated for each country using the age at death distribution presented in in Table A4. We take the mid-point *x* for each interval, calculated the corresponding *dQALY*(*x*), multiply by the proportion of deaths in that interval, and sum the products.

#### A.4 Further model inputs

The age at death distributions are provided in Table A4. Data were not available for Ireland so this was assumed to be the same as in the UK. This is supported by Omori 2020 who compared age at death distributions across Spain, Italy, and Japan.^24^ They found that although total numbers of dead varied between countries, the age distribution at death was similar.

The lifetables used in the analysis are presented in Table A5. These represent the probability of dying between the age of the lower and upper limits of each interval. Eurostat was used for Spain and Sweden but this reports only as high as age 84.^14^ After this, the UK ONS figures were used. German data from Destatis were only available to age 99 but no data were employed beyond this. ^15^

**Table A3.** Input parameters used in NHS cost-effectiveness model

| Parameter | Country | Value | Source |
| --- | --- | --- | --- |
| Date of first confirmed case (2020) | UK | 1<sup>st</sup> February | WHO coronavirus disease dashboard<sup>5</sup> |
|  | Ireland | 1<sup>st</sup> March |  |
|  | Spain | 2<sup>nd</sup> February |  |
|  | Germany | 28<sup>th</sup> January |  |
|  | Sweden | 1<sup>st</sup> February |  |
| Date of first confirmed death | UK | 7<sup>th</sup> March |  |
|  | Ireland | 12<sup>th</sup> March |  |
|  | Spain | 13<sup>th</sup> February |  |
|  | Germany | 10<sup>th</sup> March |  |
|  | Sweden | 12<sup>th</sup> March |  |
| Cases to 20<sup>th</sup> July | UK | 295372 | UK government website covid-19 cases<sup>6</sup> |
|  | Ireland | 25766 | Irish government website covid-19 cases<sup>7</sup> |
|  | Spain | 291583 | Spanish National Centre for Epidemiology website. Total over provincial level covid-19 cases<sup>8</sup> |
|  | Germany | 203500 | Tagesspiegel website covid-19 cases<sup>9</sup> |
|  | Sweden | 78135 | Public Health Agency of Sweden website.<sup>10</sup> |
| Hospitalisations (ICU and non-ICU) to 20<sup>th</sup> July | UK | 133047 | European Centre for Disease Prevention and Control hospital admissions data.<sup>11</sup> |
|  | Ireland | 1953 |  |
|  | Spain | 90897 |  |
|  | Germany | 23750 |  |
|  | Sweden | 21810 | Not reported. Calculated using hospitalisation to case ratio of 0.30 across UK, Ireland, Spain, and Germany. |
| Deaths to 20<sup>th</sup> July | UK | 41101 | UK government website covid-19 deaths<sup>6</sup> |
|  | Ireland | 1753 | Irish government website covid-19 related deaths<sup>7</sup> |
|  | Spain | 28428 | WHO coronavirus disease dashboard on covid-19 related deaths<sup>5</sup> |
|  | Germany | 9170 | Tagesspiegel website covid-19 related deaths<sup>9</sup> |
|  | Sweden | 5717 | Public Health Agency of Sweden website on covid-19 related deaths.<sup>10</sup> |
| Life expectancy | UK | Table A5 | Office of National Statistician (ONS)<sup>12</sup> |
|  | Ireland | Table A5 | Central Statistics Office (CSO)<sup>13</sup> |
|  | Spain | Table A5 | Eurostat <sup>14</sup>. UK ONS figures from age 85 to 100 as Eurostat only reports to age 84.<sup>12</sup> |
|  | Germany | Table A5 | Destatis<sup>15</sup> |
|  | Sweden | Table A5 | Eurostat <sup>14</sup>. UK ONS figures from age 85 to 100 as Eurostat only reports to age 84.<sup>12,16</sup> |
| Proportion female | All | 0.50 | Assumed for all countries and age groups to be the same |
| Age distribution at death | All | Table A4 | Various sources, details in below. |
| QALY norms per age | All | Table A6<br>**Error! Reference source not found.** | Published estimates<sup>17</sup> |
| QALYs lost per community case | All | -0.00167 | Weighted average of QALY lost per community case of flu in England over four strains<sup>18</sup> |
| QALYs lost per hospitalisation (ICU or non-ICU) | All | -0.031 | QALY loss per hospitalisation in 431 H1N1 inpatients in Spain <sup>19</sup> |
| Cost per community case | All | £0 | Assumption |
| Cost per hospitalisation (ICU or non-ICU) | All | £4,746 | National schedule of NHS costs for ICU and non-ICU, 9 day length of stay from ICNARC report on covid-19, 17% hospitalised requiring ICU. <sup>20-22</sup> |
| Duration of non-ICU hospital in CMMID | All | 8 days | CMMID model<sup>23</sup> |
| Duration of ICU stay in CMMID | All | 10 days | CMMID model<sup>23</sup> |

**Table A4.** Age distribution at death. Ireland distribution assumed same as for the UK.

| Age band (years) | UK | Sweden | Spain | Germany |
| --- | --- | --- | --- | --- |
| Source | Briggs 2020<sup>3</sup> | Swedish Public Health Authority<sup>25</sup> | Spanish Ministry of Health<sup>26</sup> | Statista data for Germany<sup>27</sup> |
| 0-9 | 4.47E-05 | 0.000174 | 0 | 0.00011 |
| 10-19 | 0.000358 | 0 | 0 | 0.00022 |
| 20-29 | 0.0017 | 0.001563 | 0.001 | 0.000989 |
| 30-39 | 0.004921 | 0.002778 | 0.003 | 0.002527 |
| 40-49 | 0.016196 | 0.007639 | 0.011 | 0.00868 |
| 50-59 | 0.052928 | 0.027951 | 0.032 | 0.03593 |
| 60-69 | 0.11476 | 0.068924 | 0.089 | 0.097572 |
| 70-79 | 0.249295 | 0.215278 | 0.241 | 0.226349 |
| 80-90 | 0.380923 | 0.415278 | 0.411 | 0.444457 |
| 90-100 | 0.178873 | 0.260417 | 0.212 | 0.183167 |

**Table A5.** Lifetables for each country. qx is probability of dying between age x and x+1.

|  | UK<br>ONS lifetables 2017-2019<sup>12</sup> |  | Ireland<br>Central Statistics Office<br>lifetables 2015-2017<sup>13</sup> |  | Spain<br>Eurostat 2018<sup>14</sup> |  | Germany<br>Destatis 2017-2019<sup>15</sup> |  | Sweden<br>Eurostat 2018<sup>14</sup> |  |
| --- | --- | --- | --- | --- | --- | --- | --- | --- | --- | --- |
| age | qx male | qx female | qx_male | qx_female | qx_male | qx_female | qx_male | qx_female | qx_male | qx_female |
| 0 | 0.004267 | 0.003536 | 0.00332355 | 0.00304020 | 0.0028 | 0.0025 | 0.0035 | 0.0029 | 0.0022 | 0.0018 |
| 1 | 0.000243 | 0.000213 | 0.00018249 | 0.00021585 | 0.0002 | 0.0002 | 0.0003 | 0.0002 | 0.0002 | 0.0003 |
| 2 | 0.000132 | 0.000127 | 0.00015815 | 0.00006683 | 0.0001 | 0.0001 | 0.0001 | 0.0001 | 0.0002 | 0.0001 |
| 3 | 0.000101 | 0.000098 | 0.00010511 | 0.00005239 | 0.0001 | 0.0001 | 0.0001 | 0.0001 | 0.0001 | 0.0001 |
| 4 | 0.000097 | 0.000068 | 0.00008877 | 0.00006425 | 0.0001 | 0.0001 | 0.0001 | 0.0001 | 0.0001 | 0.0001 |
| 5 | 0.000085 | 0.000086 | 0.00008314 | 0.00007613 | 0.0001 | 0.0001 | 0.0001 | 0.0001 | 0.0001 | 0.0001 |
| 6 | 0.000088 | 0.000082 | 0.00007538 | 0.00006452 | 0.0001 | 0.0001 | 0.0001 | 0.0001 | 0.0001 | 0.0000 |
| 7 | 0.000069 | 0.000062 | 0.00005775 | 0.00005301 | 0.0001 | 0.0001 | 0.0001 | 0.0001 | 0.0001 | 0.0001 |
| 8 | 0.000067 | 0.000064 | 0.00003262 | 0.00004637 | 0.0001 | 0.0001 | 0.0001 | 0.0001 | 0.0001 | 0.0001 |
| 9 | 0.000059 | 0.000053 | 0.00001732 | 0.00004306 | 0.0001 | 0.0000 | 0.0001 | 0.0000 | 0.0000 | 0.0000 |
| 10 | 0.000074 | 0.000064 | 0.00003031 | 0.00004232 | 0.0001 | 0.0001 | 0.0001 | 0.0001 | 0.0001 | 0.0001 |
| 11 | 0.000085 | 0.000066 | 0.00007924 | 0.00004389 | 0.0001 | 0.0001 | 0.0001 | 0.0001 | 0.0001 | 0.0001 |
| 12 | 0.000105 | 0.000060 | 0.00009941 | 0.00004787 | 0.0001 | 0.0001 | 0.0001 | 0.0001 | 0.0001 | 0.0000 |
| 13 | 0.000127 | 0.000080 | 0.00009136 | 0.00005476 | 0.0001 | 0.0001 | 0.0001 | 0.0001 | 0.0001 | 0.0001 |
| 14 | 0.000121 | 0.000101 | 0.00011522 | 0.00006548 | 0.0001 | 0.0001 | 0.0001 | 0.0001 | 0.0002 | 0.0002 |
| 15 | 0.000174 | 0.000112 | 0.00017672 | 0.00008161 | 0.0001 | 0.0001 | 0.0002 | 0.0001 | 0.0001 | 0.0001 |
| 16 | 0.000227 | 0.000147 | 0.00025543 | 0.00010570 | 0.0002 | 0.0001 | 0.0002 | 0.0001 | 0.0001 | 0.0002 |
| 17 | 0.000316 | 0.000159 | 0.00033479 | 0.00014179 | 0.0002 | 0.0002 | 0.0003 | 0.0001 | 0.0004 | 0.0002 |
| 18 | 0.000400 | 0.000227 | 0.00040443 | 0.00018852 | 0.0003 | 0.0001 | 0.0004 | 0.0002 | 0.0004 | 0.0002 |
| 19 | 0.000448 | 0.000200 | 0.00045766 | 0.00020377 | 0.0003 | 0.0001 | 0.0004 | 0.0002 | 0.0004 | 0.0003 |
| 20 | 0.000508 | 0.000190 | 0.00049306 | 0.00017518 | 0.0003 | 0.0001 | 0.0004 | 0.0002 | 0.0004 | 0.0003 |
| 21 | 0.000513 | 0.000211 | 0.00051400 | 0.00015336 | 0.0003 | 0.0002 | 0.0004 | 0.0002 | 0.0008 | 0.0003 |
| 22 | 0.000504 | 0.000229 | 0.00052696 | 0.00015172 | 0.0003 | 0.0001 | 0.0004 | 0.0002 | 0.0006 | 0.0003 |
| 23 | 0.000505 | 0.000222 | 0.00053924 | 0.00016302 | 0.0004 | 0.0001 | 0.0004 | 0.0002 | 0.0007 | 0.0004 |
| 24 | 0.000552 | 0.000222 | 0.00055336 | 0.00018190 | 0.0004 | 0.0002 | 0.0005 | 0.0002 | 0.0008 | 0.0002 |
| 25 | 0.000598 | 0.000254 | 0.00056944 | 0.00020272 | 0.0003 | 0.0002 | 0.0004 | 0.0002 | 0.0006 | 0.0002 |
| 26 | 0.000581 | 0.000262 | 0.00058759 | 0.00022370 | 0.0005 | 0.0002 | 0.0004 | 0.0002 | 0.0007 | 0.0002 |
| 27 | 0.000621 | 0.000292 | 0.00060797 | 0.00024471 | 0.0004 | 0.0001 | 0.0004 | 0.0002 | 0.0008 | 0.0004 |
| 28 | 0.000695 | 0.000314 | 0.00063074 | 0.00026569 | 0.0005 | 0.0002 | 0.0005 | 0.0002 | 0.0008 | 0.0004 |
| 29 | 0.000732 | 0.000320 | 0.00065608 | 0.00028665 | 0.0006 | 0.0002 | 0.0005 | 0.0003 | 0.0006 | 0.0003 |
| 30 | 0.000771 | 0.000370 | 0.00068420 | 0.00030768 | 0.0005 | 0.0003 | 0.0005 | 0.0003 | 0.0007 | 0.0004 |
| 31 | 0.000835 | 0.000394 | 0.00071535 | 0.00032895 | 0.0005 | 0.0002 | 0.0006 | 0.0003 | 0.0006 | 0.0003 |
| 32 | 0.000835 | 0.000453 | 0.00074979 | 0.00035072 | 0.0005 | 0.0002 | 0.0006 | 0.0003 | 0.0006 | 0.0005 |
| 33 | 0.000929 | 0.000486 | 0.00078782 | 0.00037335 | 0.0005 | 0.0003 | 0.0007 | 0.0004 | 0.0008 | 0.0004 |
| 34 | 0.000957 | 0.000556 | 0.00082979 | 0.00039728 | 0.0006 | 0.0003 | 0.0007 | 0.0004 | 0.0009 | 0.0004 |
| 35 | 0.001073 | 0.000582 | 0.00087609 | 0.00042310 | 0.0006 | 0.0003 | 0.0008 | 0.0004 | 0.0008 | 0.0004 |
| 36 | 0.001146 | 0.000645 | 0.00092712 | 0.00045150 | 0.0007 | 0.0003 | 0.0009 | 0.0005 | 0.0008 | 0.0003 |
| 37 | 0.001309 | 0.000784 | 0.00098340 | 0.00048334 | 0.0007 | 0.0004 | 0.0010 | 0.0005 | 0.0008 | 0.0004 |
| 38 | 0.001261 | 0.000732 | 0.00104544 | 0.00051970 | 0.0007 | 0.0004 | 0.0010 | 0.0006 | 0.0008 | 0.0004 |
| 39 | 0.001399 | 0.000852 | 0.00111386 | 0.00056191 | 0.0008 | 0.0004 | 0.0011 | 0.0006 | 0.0009 | 0.0004 |
| 40 | 0.001529 | 0.000902 | 0.00118935 | 0.00061115 | 0.0009 | 0.0005 | 0.0012 | 0.0006 | 0.0011 | 0.0005 |
| 41 | 0.001670 | 0.000986 | 0.00127267 | 0.00066827 | 0.0010 | 0.0006 | 0.0014 | 0.0007 | 0.0009 | 0.0007 |
| 42 | 0.001818 | 0.001073 | 0.00136468 | 0.00073417 | 0.0011 | 0.0006 | 0.0014 | 0.0008 | 0.0011 | 0.0006 |
| 43 | 0.002002 | 0.001171 | 0.00146634 | 0.00080983 | 0.0012 | 0.0007 | 0.0016 | 0.0009 | 0.0010 | 0.0008 |
| 44 | 0.002093 | 0.001316 | 0.00157875 | 0.00089631 | 0.0012 | 0.0007 | 0.0017 | 0.0009 | 0.0013 | 0.0006 |
| 45 | 0.002347 | 0.001425 | 0.00170313 | 0.00099473 | 0.0014 | 0.0009 | 0.0019 | 0.0011 | 0.0011 | 0.0008 |
| 46 | 0.002488 | 0.001564 | 0.00184085 | 0.00110627 | 0.0017 | 0.0010 | 0.0021 | 0.0012 | 0.0015 | 0.0009 |
| 47 | 0.002696 | 0.001695 | 0.00199346 | 0.00123207 | 0.0019 | 0.0011 | 0.0024 | 0.0013 | 0.0015 | 0.0011 |
| 48 | 0.002852 | 0.001840 | 0.00216271 | 0.00137323 | 0.0023 | 0.0011 | 0.0026 | 0.0015 | 0.0015 | 0.0011 |
| 49 | 0.003189 | 0.001943 | 0.00235057 | 0.00153076 | 0.0024 | 0.0014 | 0.0029 | 0.0017 | 0.0019 | 0.0013 |
| 50 | 0.003379 | 0.002169 | 0.00255924 | 0.00170545 | 0.0029 | 0.0015 | 0.0032 | 0.0018 | 0.0022 | 0.0013 |
| 51 | 0.003606 | 0.002358 | 0.00279124 | 0.00189784 | 0.0033 | 0.0016 | 0.0036 | 0.0021 | 0.0021 | 0.0015 |
| 52 | 0.003907 | 0.002557 | 0.00304937 | 0.00210805 | 0.0035 | 0.0017 | 0.0040 | 0.0023 | 0.0029 | 0.0017 |
| 53 | 0.004125 | 0.002697 | 0.00333683 | 0.00233599 | 0.0039 | 0.0021 | 0.0046 | 0.0025 | 0.0032 | 0.0018 |
| 54 | 0.004478 | 0.002914 | 0.00365720 | 0.00258262 | 0.0047 | 0.0022 | 0.0051 | 0.0028 | 0.0031 | 0.0020 |
| 55 | 0.004760 | 0.003194 | 0.00401454 | 0.00284955 | 0.0052 | 0.0023 | 0.0057 | 0.0031 | 0.0034 | 0.0023 |
| 56 | 0.005389 | 0.003542 | 0.00441345 | 0.00313865 | 0.0056 | 0.0026 | 0.0064 | 0.0035 | 0.0039 | 0.0025 |
| 57 | 0.005856 | 0.003816 | 0.00485909 | 0.00345214 | 0.0060 | 0.0029 | 0.0072 | 0.0038 | 0.0042 | 0.0030 |
| 58 | 0.006394 | 0.004230 | 0.00535733 | 0.00379261 | 0.0067 | 0.0031 | 0.0078 | 0.0042 | 0.0046 | 0.0035 |
| 59 | 0.006930 | 0.004616 | 0.00591479 | 0.00416312 | 0.0072 | 0.0034 | 0.0088 | 0.0046 | 0.0056 | 0.0034 |
| 60 | 0.007595 | 0.005054 | 0.00653895 | 0.00456726 | 0.0080 | 0.0036 | 0.0096 | 0.0052 | 0.0063 | 0.0041 |
| 61 | 0.008313 | 0.005492 | 0.00723827 | 0.00500927 | 0.0089 | 0.0038 | 0.0107 | 0.0057 | 0.0067 | 0.0047 |
| 62 | 0.009223 | 0.006264 | 0.00802233 | 0.00549415 | 0.0097 | 0.0042 | 0.0118 | 0.0061 | 0.0077 | 0.0051 |
| 63 | 0.010178 | 0.006714 | 0.00890193 | 0.00602782 | 0.0105 | 0.0045 | 0.0129 | 0.0068 | 0.0081 | 0.0058 |
| 64 | 0.010947 | 0.007264 | 0.00988932 | 0.00661725 | 0.0111 | 0.0050 | 0.0141 | 0.0074 | 0.0095 | 0.0057 |
| 65 | 0.012025 | 0.007988 | 0.01099831 | 0.00727075 | 0.0121 | 0.0050 | 0.0153 | 0.0079 | 0.0107 | 0.0067 |
| 66 | 0.013330 | 0.008573 | 0.01224452 | 0.00799812 | 0.0131 | 0.0057 | 0.0167 | 0.0088 | 0.0117 | 0.0075 |
| 67 | 0.014437 | 0.009380 | 0.01364559 | 0.00881107 | 0.0143 | 0.0062 | 0.0179 | 0.0094 | 0.0129 | 0.0081 |
| 68 | 0.015740 | 0.010330 | 0.01522143 | 0.00972357 | 0.0150 | 0.0062 | 0.0194 | 0.0103 | 0.0136 | 0.0093 |
| 69 | 0.017289 | 0.011296 | 0.01699450 | 0.01075229 | 0.0169 | 0.0072 | 0.0207 | 0.0114 | 0.0153 | 0.0108 |
| 70 | 0.018286 | 0.012437 | 0.01899014 | 0.01191729 | 0.0190 | 0.0078 | 0.0226 | 0.0127 | 0.0168 | 0.0114 |
| 71 | 0.020276 | 0.013337 | 0.02123685 | 0.01324270 | 0.0197 | 0.0085 | 0.0243 | 0.0136 | 0.0181 | 0.0132 |
| 72 | 0.022326 | 0.015241 | 0.02376674 | 0.01475770 | 0.0219 | 0.0093 | 0.0264 | 0.0151 | 0.0208 | 0.0138 |
| 73 | 0.025500 | 0.017347 | 0.02661587 | 0.01649767 | 0.0235 | 0.0105 | 0.0284 | 0.0165 | 0.0209 | 0.0159 |
| 74 | 0.028123 | 0.019167 | 0.02982471 | 0.01850572 | 0.0260 | 0.0123 | 0.0312 | 0.0181 | 0.0247 | 0.0177 |
| 75 | 0.031402 | 0.021437 | 0.03343860 | 0.02083453 | 0.0284 | 0.0140 | 0.0339 | 0.0199 | 0.0276 | 0.0197 |
| 76 | 0.035115 | 0.024217 | 0.03750824 | 0.02354813 | 0.0303 | 0.0156 | 0.0367 | 0.0215 | 0.0320 | 0.0205 |
| 77 | 0.038838 | 0.027308 | 0.04209018 | 0.02671326 | 0.0344 | 0.0175 | 0.0405 | 0.0239 | 0.0350 | 0.0230 |
| 78 | 0.043521 | 0.030961 | 0.04724736 | 0.03039745 | 0.0408 | 0.0213 | 0.0446 | 0.0270 | 0.0383 | 0.0278 |
| 79 | 0.048099 | 0.034481 | 0.05304958 | 0.03467553 | 0.0441 | 0.0245 | 0.0499 | 0.0310 | 0.0445 | 0.0310 |
| 80 | 0.053982 | 0.038463 | 0.05957403 | 0.03962946 | 0.0504 | 0.0289 | 0.0554 | 0.0355 | 0.0497 | 0.0335 |
| 81 | 0.060071 | 0.043634 | 0.06690568 | 0.04534759 | 0.0557 | 0.0332 | 0.0625 | 0.0412 | 0.0590 | 0.0406 |
| 82 | 0.066512 | 0.048958 | 0.07513766 | 0.05192313 | 0.0626 | 0.0377 | 0.0709 | 0.0485 | 0.0660 | 0.0462 |
| 83 | 0.075396 | 0.056269 | 0.08437146 | 0.05945201 | 0.0699 | 0.0436 | 0.0794 | 0.0556 | 0.0757 | 0.0516 |
| 84 | 0.084755 | 0.063935 | 0.09471695 | 0.06802964 | 0.0811 | 0.0514 | 0.0907 | 0.0645 | 0.0833 | 0.0619 |
| 85 | 0.094664 | 0.072462 | 0.10629219 | 0.07774677 |  |  | 0.1019 | 0.0742 |  |  |
| 86 | 0.106853 | 0.083094 | 0.11922282 | 0.08868416 |  |  | 0.1160 | 0.0861 |  |  |
| 87 | 0.118585 | 0.093462 | 0.13364105 | 0.10090621 |  |  | 0.1292 | 0.0990 |  |  |
| 88 | 0.133359 | 0.106434 | 0.14968404 | 0.11445357 |  |  | 0.1474 | 0.1137 |  |  |
| 89 | 0.149851 | 0.118942 | 0.16749159 | 0.12933523 |  |  | 0.1640 | 0.1288 |  |  |
| 90 | 0.159530 | 0.134387 | 0.18720261 | 0.14552025 |  |  | 0.1821 | 0.1472 |  |  |
| 91 | 0.179055 | 0.150755 | 0.20830632 | 0.16293008 |  |  | 0.1978 | 0.1659 |  |  |
| 92 | 0.196950 | 0.167079 | 0.22776219 | 0.18143198 |  |  | 0.2239 | 0.1880 |  |  |
| 93 | 0.215044 | 0.184344 | 0.24267951 | 0.20083448 |  |  | 0.2427 | 0.2093 |  |  |
| 94 | 0.238086 | 0.204467 | 0.25301883 | 0.22088580 |  |  | 0.2675 | 0.2284 |  |  |
| 95 | 0.261012 | 0.228210 | 0.25944618 | 0.24127563 |  |  | 0.2877 | 0.2539 |  |  |
| 96 | 0.286714 | 0.250765 | 0.26290017 | 0.26164101 |  |  | 0.3149 | 0.2781 |  |  |
| 97 | 0.304113 | 0.267058 | 0.26447343 | 0.28157596 |  |  | 0.3418 | 0.2941 |  |  |
| 98 | 0.325892 | 0.291260 | 0.26532494 | 0.30064477 |  |  | 0.3584 | 0.3175 |  |  |
| 99 | 0.369540 | 0.309526 | 0.26663580 | 0.31839759 |  |  | 0.3816 | 0.3408 |  |  |
| 100 | 0.384386 | 0.343363 | 0.26960804 | 0.33438743 |  |  |  |  |  |  |

**Table A6.** QALY norms for each country by age band. ^17^

| Age band | UK | Ireland | Spain | Germany | Sweden |
| --- | --- | --- | --- | --- | --- |
| 0-17 | 1 | 1 | 1 | 1 | 1 |
| 18-24 | 0.94 | 0.94 | 0.982 | 0.972 | 0.888 |
| 25-34 | 0.927 | 0.927 | 0.975 | 0.973 | 0.893 |
| 35-44 | 0.911 | 0.911 | 0.949 | 0.966 | 0.868 |
| 45-54 | 0.847 | 0.847 | 0.923 | 0.945 | 0.835 |
| 55-64 | 0.799 | 0.799 | 0.901 | 0.922 | 0.813 |
| 65-74 | 0.779 | 0.779 | 0.891 | 0.891 | 0.836 |
| 75+ | 0.726 | 0.726 | 0.781 | 0.839 | 0.701 |

## Appendix B. Country-level response details

**Table A7.**
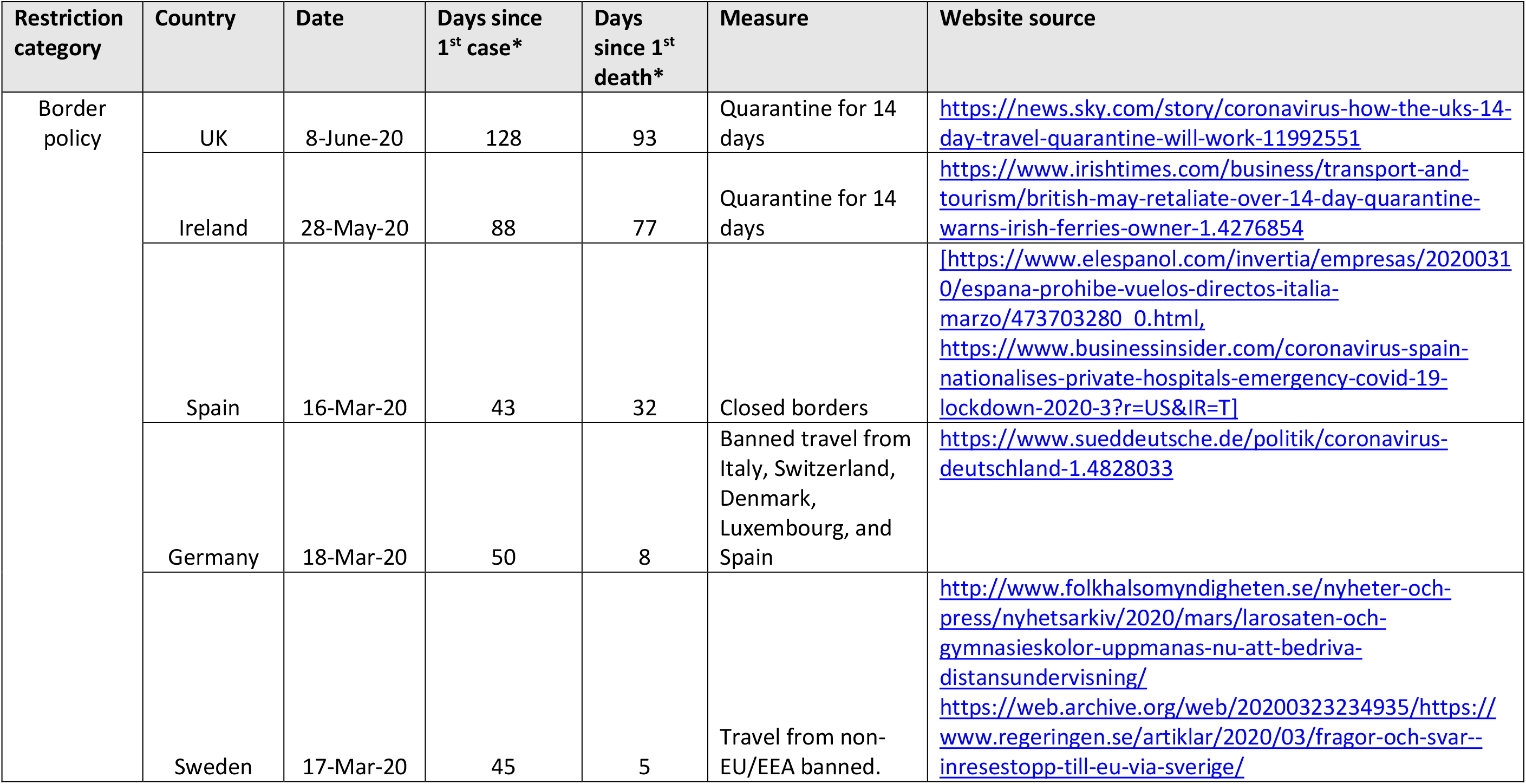
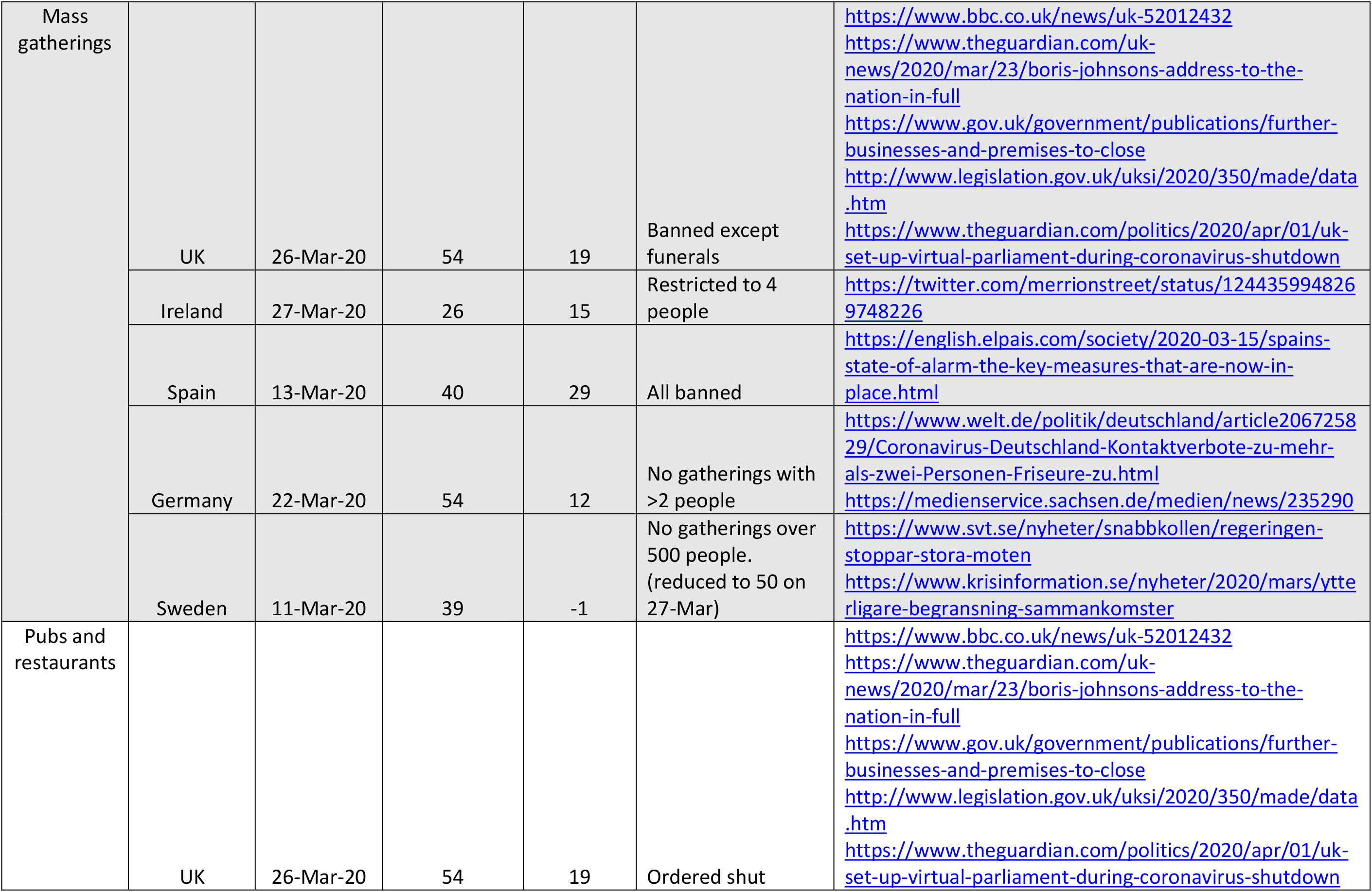
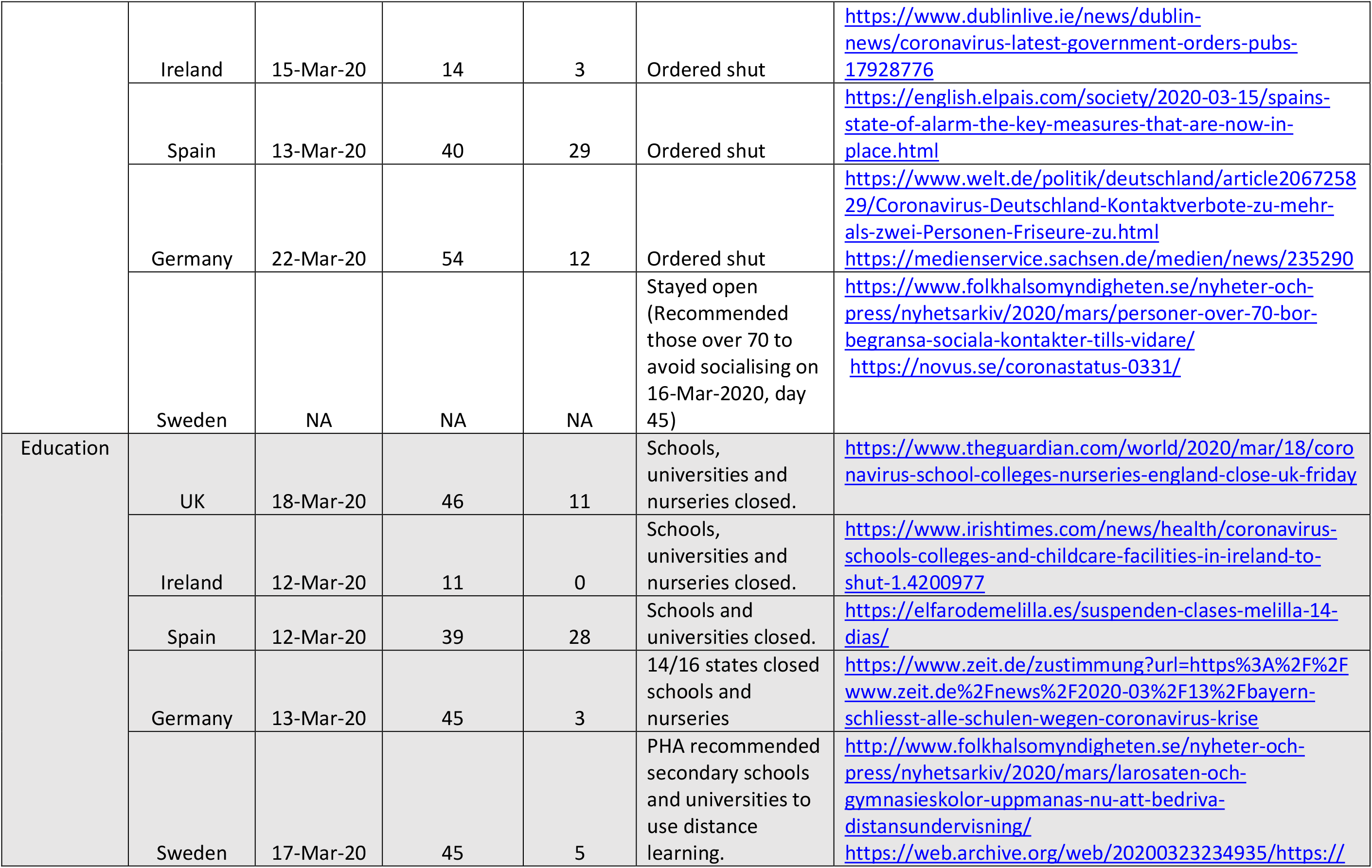
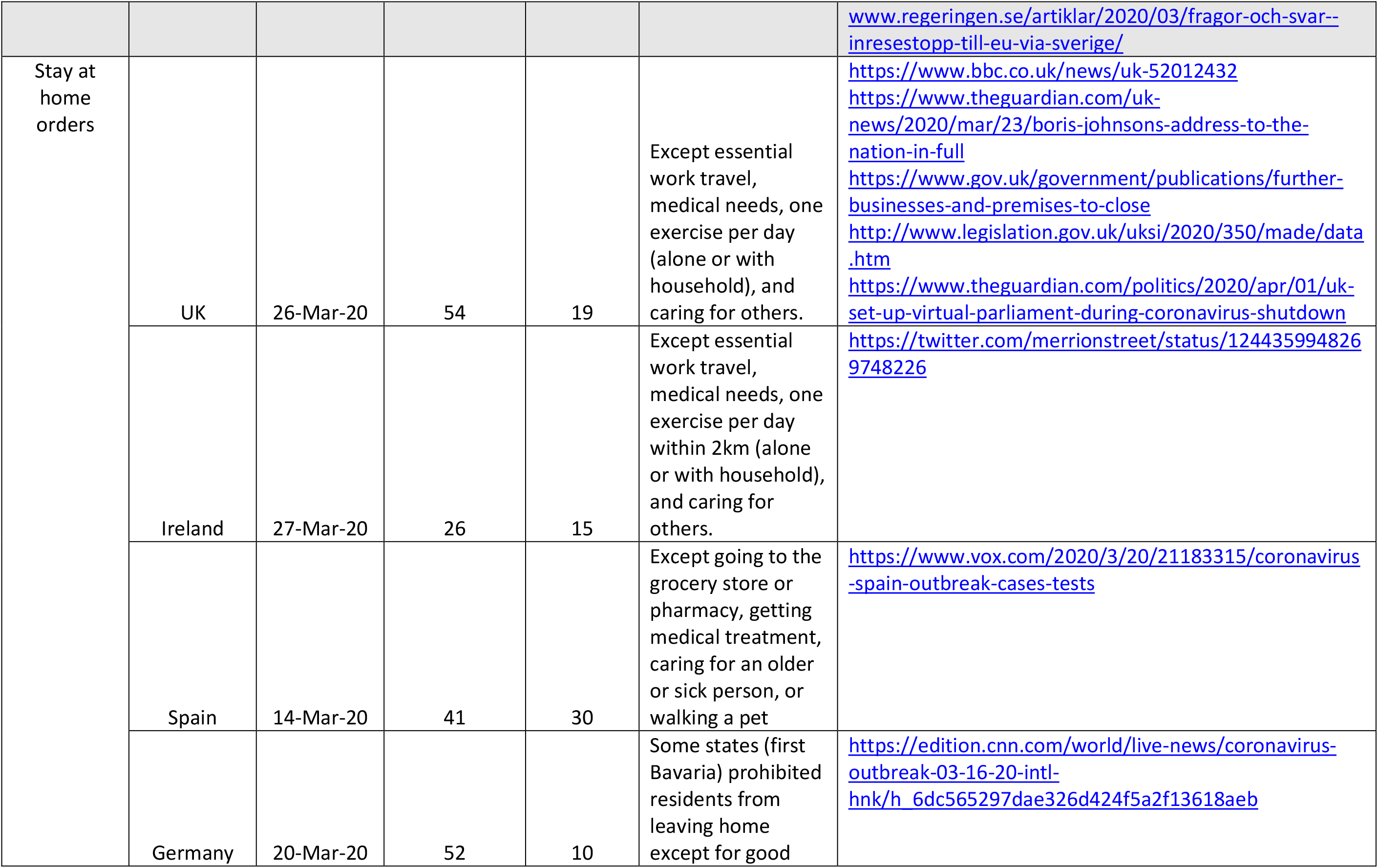
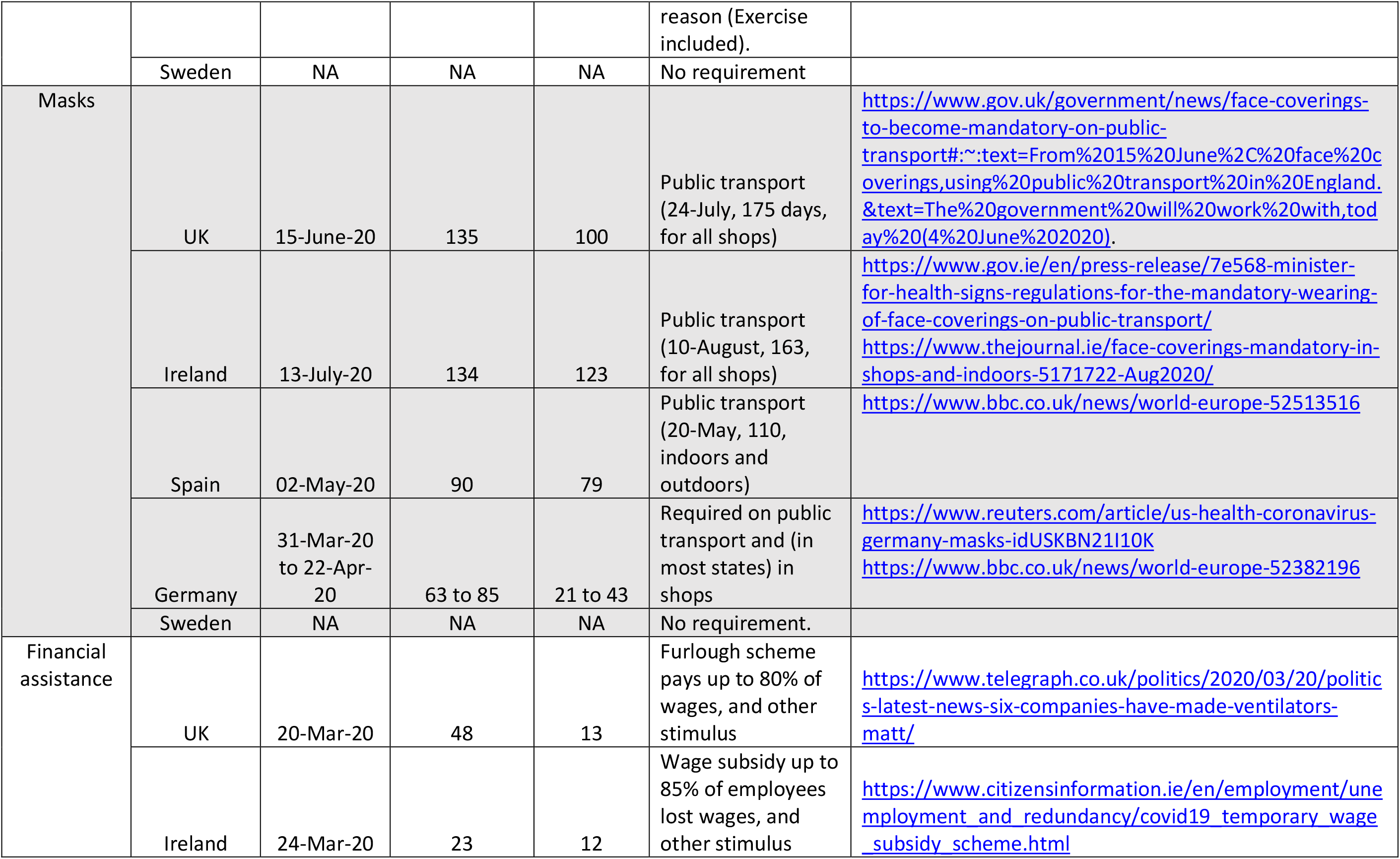
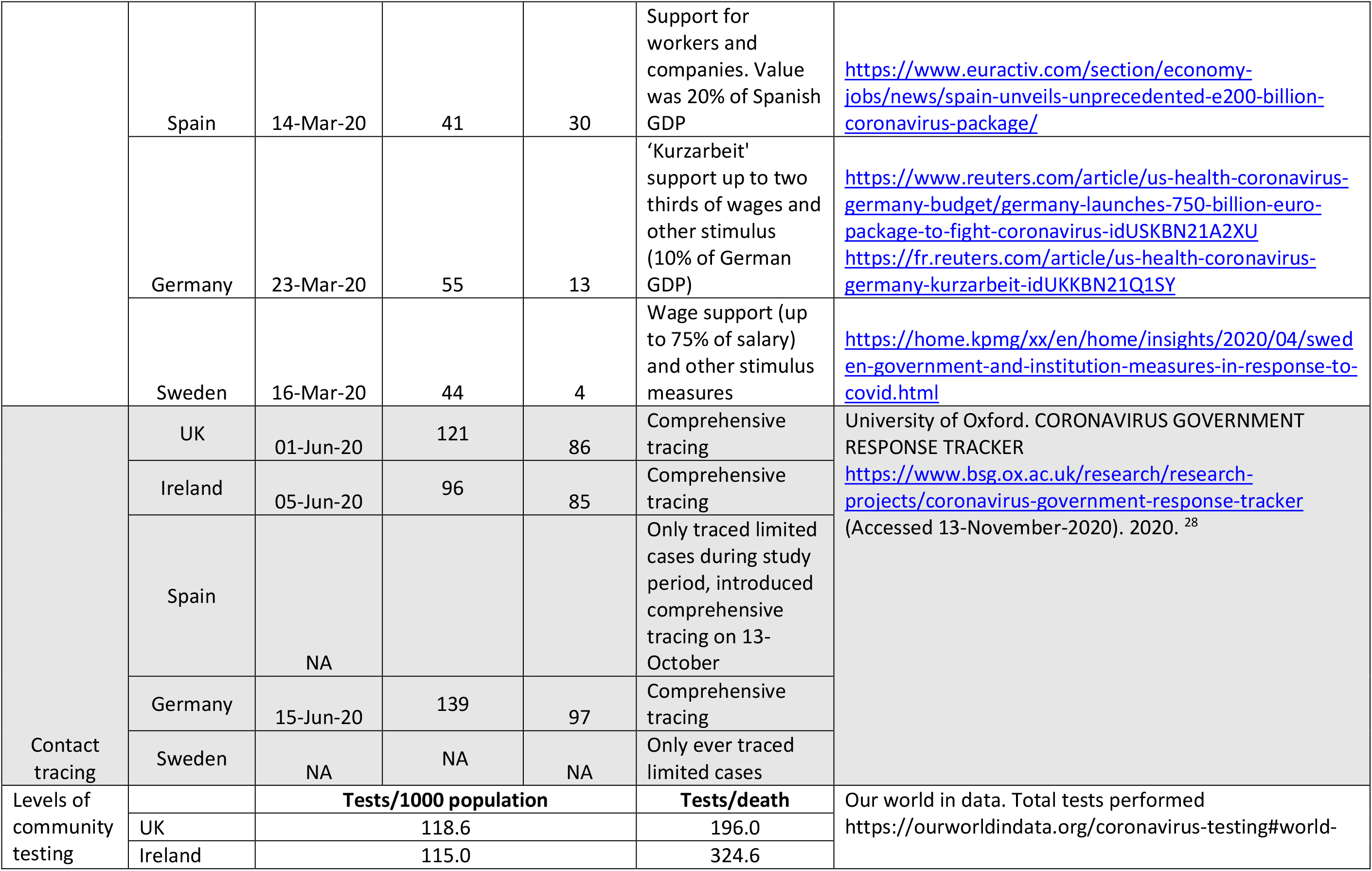
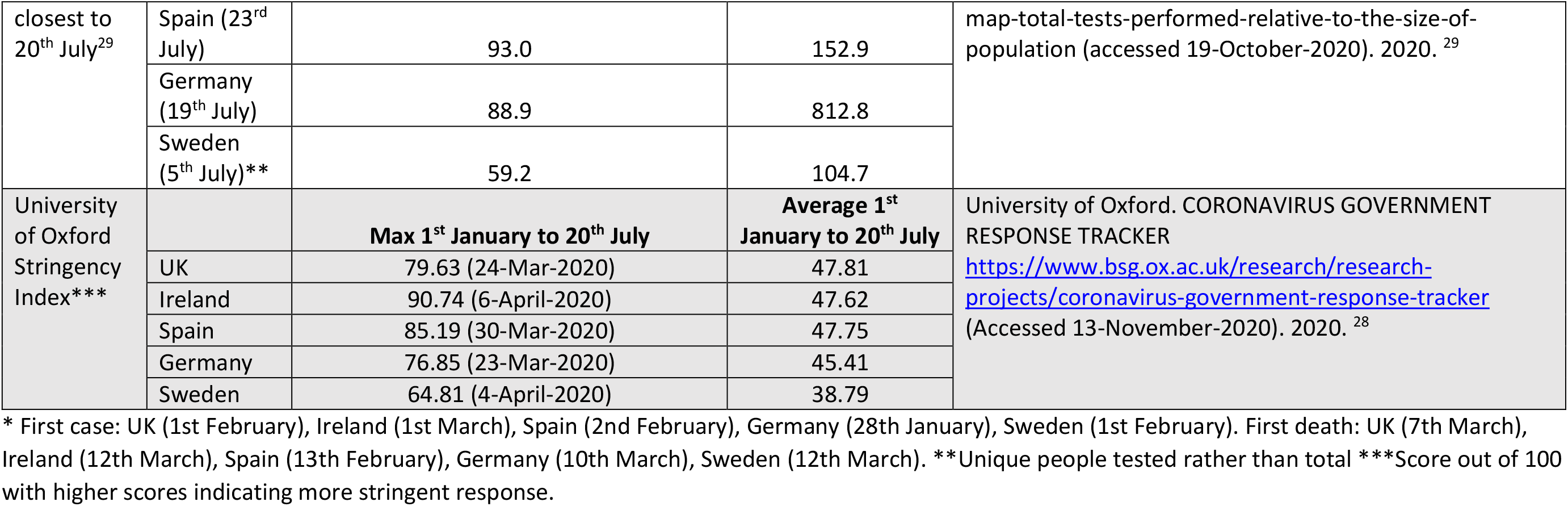
Description and timing, relative to 1^st^ case and 1^st^ death in each country, of country responses to covid-19

## Appendix C. Costing details

The cost of hospitalisation was calculated using ICU and non-ICU costs, the length of stay for a hospitalisation, and the proportion hospital admissions requiring ICU. We used the National schedule of NHS costs to calculate a hospital stay cost of £2161 as a weighted average of “Unspecified Acute Lower Respiratory Infection with Interventions”.^21^ We also used this source to calculate an ICU daily cost of £1504 as a weighted average of “Non-specific, general adult critical care patients predominate”). A study of 20,133 patients hospitalised by covid-19 in the UK reported that 17% of hospitalised patients required admission to high dependency or ICU.^20^ The Intensive Care National Audit and Research Centre (ICNARC) report on covid-19 in critical care published on 24 July 2020 gives median critical care length of stay in patients who died in critical care (9 days, IQR 5-16, in 4078 patients), were discharged from critical care but died in hospital (4 days, IQR 2-11, in 289 patients, or were discharged from hospital (12 days, IQR 5-27, in 5344 patients).^22^ Using a sample size weighted average, we get a median length of stay in critical care of 10.50 days.

## Appendix C. Further results

**Table A8.** Observed and simulated total and per capita cases, hospitalisations, and deaths due to covid-19 to 20^th^ July 2020. Also reported are estimated QALYs lost per death for SMR=1.1. Absolute values are presented for the observed “mitigation” scenarios and incremental results for modelled “no mitigation” scenarios. These differences represent cases, hospitalisations, and deaths prevented by government intervention.

| Scenario | Country | QALYs lost per death | Cases | Hospitalisations | Deaths | Cases per capita | Hospitalisations per capita | Deaths per capita |
| --- | --- | --- | --- | --- | --- | --- | --- | --- |
| Observed<br>“Mitigation” | UK | 5.77 | 295,372 | 133,047 | 41,101 | 0.0043 | 0.0020 | 0.0006 |
|  | Ireland | 5.66 | 25,766 | 1,953 | 1,753 | 0.0052 | 0.0004 | 0.0004 |
|  | Spain | 5.77 | 291,583 | 90,897 | 28,428 | 0.0062 | 0.0019 | 0.0006 |
|  | Germany | 5.89 | 203,500 | 23,750 | 9,170 | 0.0024 | 0.0003 | 0.0001 |
|  | Sweden | 4.77 | 78,135 | 21,810* | 5,717 | 0.0077 | 0.0022 | 0.0006 |
|  | Country | QALYs lost per death | Cases prevented | Hospitalisations prevented | Deaths prevented | Cases prevented per capita | Hospitalisations prevented per capita | Deaths prevented per capita |
| A<br>$R_0 = 2.7$<br>“No mitigation” | UK | 8.72 | 29,297,252 | 2,269,469 | 391,003 | 0.4312 | 0.0334 | 0.0058 |
|  | Ireland | 9.39 | 1,938,838 | 113,784 | 19,604 | 0.3919 | 0.0230 | 0.0040 |
|  | Spain | 9.59 | 19,439,206 | 1,640,881 | 282,719 | 0.4157 | 0.0351 | 0.0060 |
|  | Germany | 9.73 | 34,764,563 | 2,928,507 | 504,445 | 0.4147 | 0.0349 | 0.0060 |
|  | Sweden | 8.94 | 4,099,594 | 330,298 | 56,906 | 0.4055 | 0.0327 | 0.0056 |
| B<br>All $R_0 = 1.6$<br>“No mitigation” | UK | 9.39 | 13,670,017 | 625,276 | 71,045 | 0.2012 | 0.0092 | 0.0010 |
|  | Ireland | 10.06 | 1,174,784 | 56,130 | 8,942 | 0.2374 | 0.0113 | 0.0018 |
|  | Spain | 10.29 | 10,068,073 | 570,549 | 73,269 | 0.2153 | 0.0122 | 0.0016 |
|  | Germany | 10.75 | 13,015,024 | 550,401 | 58,509 | 0.1553 | 0.0066 | 0.0007 |
|  | Sweden | 9.67 | 2,382,932 | 144,735 | 20,734 | 0.2357 | 0.0143 | 0.0021 |
| C<br>All $R_0 = 3.9$<br>“No mitigation” | UK | 8.54 | 32,247,215 | 2,630,081 | 453,159 | 0.4746 | 0.0387 | 0.0067 |
|  | Ireland | 9.11 | 2,193,083 | 138,433 | 23,852 | 0.4432 | 0.0280 | 0.0048 |
|  | Spain | 9.40 | 22,152,800 | 1,972,247 | 339,815 | 0.4738 | 0.0422 | 0.0073 |
|  | Germany | 9.42 | 39,533,443 | 3,574,784 | 615,930 | 0.4716 | 0.0426 | 0.0073 |
|  | Sweden | 8.68 | 4,644,719 | 397,134 | 68,426 | 0.4594 | 0.0393 | 0.0068 |

\*Total ICU and non-ICU hospitalisation data not available for Sweden. Used average hospitalisations per cases of other countries (0.279) to impute.

**Table A9.** Healthcare impacts for SMR=1.1 scenarios broken down by impact from cases, hospitalisations, and deaths. Comparisons up to 20^th^ July 2020. (INB = Incremental net health benefit, PP = Per capita).

|  |  | Cases only |  |  |  | Hospitalisation only |  |  |  | Death only |  |  |  |
| --- | --- | --- | --- | --- | --- | --- | --- | --- | --- | --- | --- | --- | --- |
| Scenario | Country | Incremental QALYs (millions) | Incremental costs (£Billions) | INB PP at £20,000/QALY (£) | INB PP at £30,000/QALY (£) | Incremental QALYs (millions) | Incremental costs (£Billions) | INB PP at £20,000/QALY (£) | INB PP at £30,000/QALY (£) | Incremental QALYs (millions) | Incremental costs (£Billions) | INB PP at £20,000/QALY (£) | INB PP at £30,000/QALY (£) |
| A<br>$R_0 = 2.7$ | UK | 0.05 | 0.00 | 14.27 | 21.40 | 0.07 | -32.94 | 504.26 | 514.01 | 3.05 | 0.00 | 898.04 | 1,347.06 |
|  | Ireland | 0.00 | 0.00 | 12.92 | 19.39 | 0.00 | -1.67 | 352.42 | 359.43 | 0.16 | 0.00 | 629.22 | 943.83 |
|  | Spain | 0.03 | 0.00 | 13.69 | 20.53 | 0.05 | -23.84 | 530.46 | 540.73 | 2.22 | 0.00 | 948.45 | 1,422.68 |
|  | Germany | 0.06 | 0.00 | 13.78 | 20.67 | 0.09 | -43.22 | 537.07 | 547.81 | 4.32 | 0.00 | 1,030.33 | 1,545.50 |
|  | Sweden | 0.01 | 0.00 | 13.29 | 19.94 | 0.01 | -4.78 | 491.91 | 501.37 | 0.45 | 0.00 | 882.97 | 1,324.45 |
| B<br>$R_0 = 1.6$ | UK | 0.02 | 0.00 | 6.58 | 9.87 | 0.02 | -8.61 | 131.18 | 133.42 | 0.28 | 0.00 | 82.74 | 124.10 |
|  | Ireland | 0.00 | 0.00 | 7.76 | 11.64 | 0.00 | -0.82 | 172.76 | 176.15 | 0.07 | 0.00 | 272.78 | 409.17 |
|  | Spain | 0.02 | 0.00 | 6.99 | 10.48 | 0.01 | -8.00 | 177.51 | 180.69 | 0.42 | 0.00 | 180.05 | 270.08 |
|  | Germany | 0.02 | 0.00 | 5.11 | 7.66 | 0.02 | -8.03 | 99.68 | 101.63 | 0.46 | 0.00 | 110.50 | 165.75 |
|  | Sweden | 0.00 | 0.00 | 7.62 | 11.43 | 0.00 | -2.04 | 208.93 | 212.70 | 0.14 | 0.00 | 278.87 | 418.30 |
| C<br>$R_0 = 3.9$ | UK | 0.05 | 0.00 | 15.72 | 23.58 | 0.08 | -38.28 | 586.09 | 597.48 | 3.52 | 0.00 | 1,035.55 | 1,553.33 |
|  | Ireland | 0.00 | 0.00 | 14.64 | 21.96 | 0.00 | -2.04 | 429.23 | 437.78 | 0.19 | 0.00 | 762.71 | 1,144.07 |
|  | Spain | 0.04 | 0.00 | 15.63 | 23.44 | 0.06 | -28.75 | 639.73 | 652.20 | 2.66 | 0.00 | 1,137.23 | 1,705.84 |
|  | Germ<br>any | 0.07 | 0.00 | 15.68 | 23.52 | 0.11 | -52.79 | 655.93 | 669.06 | 5.18 | 0.00 | 1,235.98 | 1,853.97 |
|  | Swed<br>en | 0.01 | 0.00 | 15.10 | 22.65 | 0.01 | -5.77 | 593.83 | 605.34 | 0.54 | 0.00 | 1,059.14 | 1,588.71 |

**Table A10.**
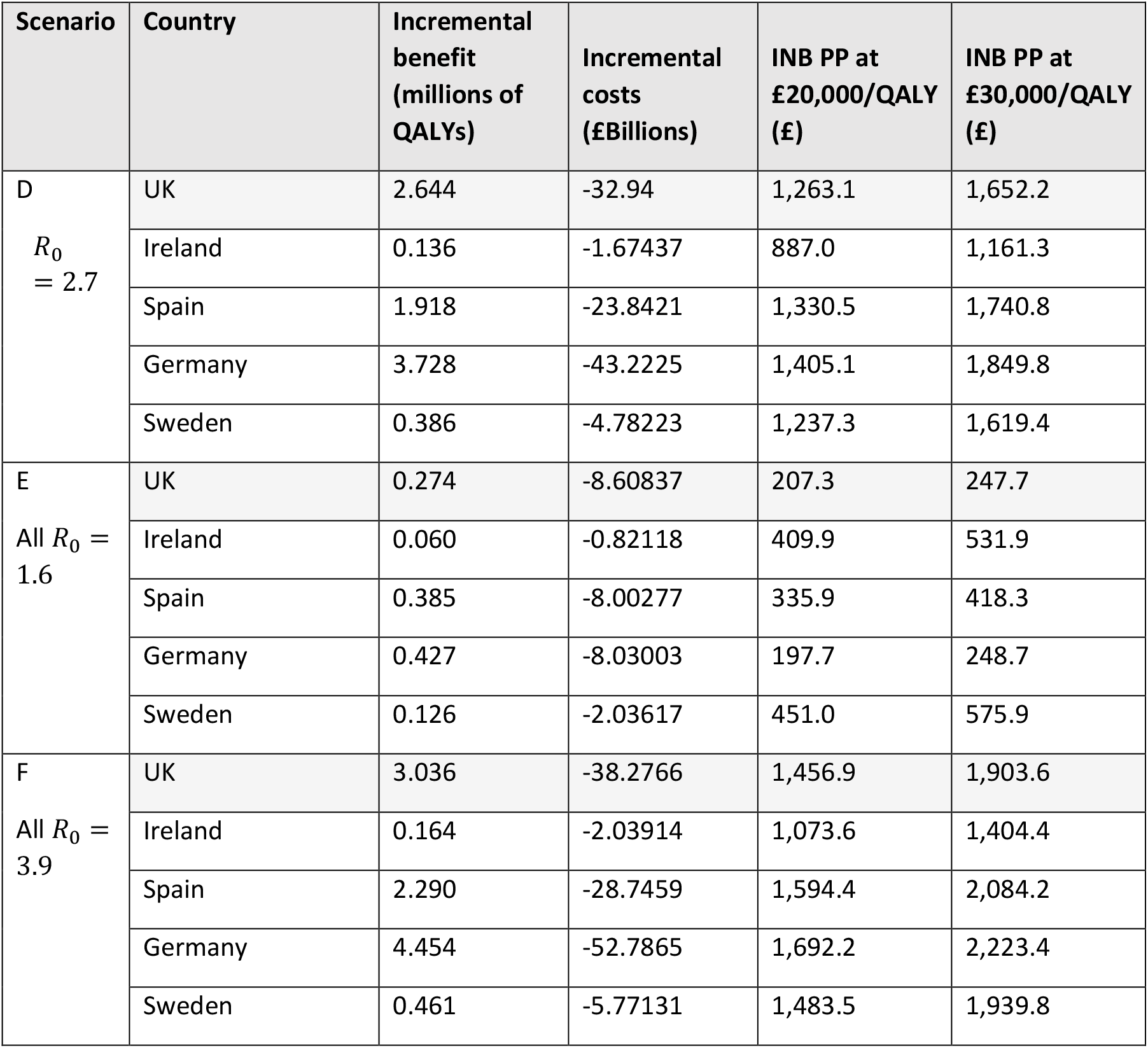
Healthcare benefits, costs, and net benefits of the government response for SMR=2.0 scenarios. Scenarios are the “No mitigation” simulations from the CMMID model. Comparisons up to 20^th^ July 2020. (INB = Incremental net health benefit, PP = Per capita).

| Scenario | Country | Incremental benefit (millions of QALYs) | Incremental costs (£Billions) | INB PP at £20,000/QALY (£) | INB PP at £30,000/QALY (£) |
| --- | --- | --- | --- | --- | --- |
| D<br>$R_0 = 2.7$ | UK | 2.644 | -32.94 | 1,263.1 | 1,652.2 |
|  | Ireland | 0.136 | -1.67437 | 887.0 | 1,161.3 |
|  | Spain | 1.918 | -23.8421 | 1,330.5 | 1,740.8 |
|  | Germany | 3.728 | -43.2225 | 1,405.1 | 1,849.8 |
|  | Sweden | 0.386 | -4.78223 | 1,237.3 | 1,619.4 |
| E<br>All $R_0 = 1.6$ | UK | 0.274 | -8.60837 | 207.3 | 247.7 |
|  | Ireland | 0.060 | -0.82118 | 409.9 | 531.9 |
|  | Spain | 0.385 | -8.00277 | 335.9 | 418.3 |
|  | Germany | 0.427 | -8.03003 | 197.7 | 248.7 |
|  | Sweden | 0.126 | -2.03617 | 451.0 | 575.9 |
| F<br>All $R_0 = 3.9$ | UK | 3.036 | -38.2766 | 1,456.9 | 1,903.6 |
|  | Ireland | 0.164 | -2.03914 | 1,073.6 | 1,404.4 |
|  | Spain | 2.290 | -28.7459 | 1,594.4 | 2,084.2 |
|  | Germany | 4.454 | -52.7865 | 1,692.2 | 2,223.4 |
|  | Sweden | 0.461 | -5.77131 | 1,483.5 | 1,939.8 |

**Table A11.** Incremental net health benefit (INB) at £/QALY thresholds justified by NICE end-of-life criteria (£50,000), societal perspective (£10,000 and £70,000), and UK Cancer drugs fund (£220,000).

| Scenario | Country | GDP los<br>PP (£)<br>Scenario:<br>Sweden<br>growth<br>loss as no<br>mitigatio<br>n | GDP los<br>PP (£)<br>Scenario:<br>All GDP<br>reductio<br>n due to<br>response | INB PP at<br>£10,000/QAL<br>Y PP<br>(£) | INB PP at<br>£20,000/QAL<br>Y PP<br>(£) | INB PP at<br>£30,000/QAL<br>Y<br>(£) | INB PP at<br>£50,000/QAL<br>Y<br>(£) | INB PP at<br>£70,000/QAL<br>Y<br>(£) | INB PP at<br>£220,000/QAL<br>Y<br>(£) |
| --- | --- | --- | --- | --- | --- | --- | --- | --- | --- |
| A<br>$R_0 = 2.7$,<br>SMR=1.1<br>Base case | UK | 1,455.17 | 3,075.07 | 951 | 1,417 | 1,883 | 2,814 | 3,746 | 10,735 |
|  | Ireland | 211.62 | 3,332.97 | 665 | 995 | 1,323 | 1,975 | 2,629 | 7,541 |
|  | Spain | 1,727.98 | 2,927.40 | 1,001 | 1,493 | 1,984 | 2,966 | 3,949 | 11,317 |
|  | German<br>y | 376.05 | 2,224.95 | 1,048 | 1,581 | 2,114 | 3,180 | 4,246 | 12,238 |
|  | Sweden | 0 | 2,098.85 | 931 | 1,388 | 1,846 | 2,762 | 3,678 | 10,547 |
| E<br>$R_0 = 1.6$,<br>SMR=2.0<br>Most<br>unfavourabl<br>e | UK | 1,455.17 | 3,075.07 | 167 | 207.3 | 247.7 | 328 | 409 | 1,014 |
|  | Ireland | 211.62 | 3,332.97 | 287 | 409.9 | 531.9 | 772 | 1,015 | 2,834 |
|  | Spain | 1,727.98 | 2,927.40 | 253 | 335.9 | 418.3 | 583 | 748 | 1,983 |
|  | German<br>y | 376.05 | 2,224.95 | 147 | 197.7 | 248.7 | 350 | 452 | 1,216 |
|  | Sweden | 0 | 2,098.85 | 326 | 451 | 575.9 | 824 | 1,074 | 2,943 |

## References

1. WHO. WHO Coronavirus Disease (COVID-19) Dashboard https://covid19.who.int/region/euro/country/ (Accessed 13-November-2020). 2020.

2. IMF. World Economic Outlook Database, October 2019. 15 October 2019. https://www.imf.org/external/pubs/ft/weo/2019/02/weodata/index.aspx (Accessed 15-September-2020). 2019.

3. IMF. World Economic Outlook, January 2020: Tentative Stabilization, Sluggish Recovery? https://www.imf.org/en/Publications/WEO/Issues/2020/01/20/weo-update-january2020 (Accessed 1-August-2020). 2020.

4. Scally G, Jacobson B, Abbasi K. The UK’s public health response to covid-19. BMJ 2020; 369: m1932.

5. Rowthorn R, Maciejowski J. A cost-benefit analysis of the COVID-19 disease. Oxford Review of Economic Policy 2020.

6. Miles D, Stedman M, Heald AH. “Stay at Home, Protect the National Health Service, Save Lives”: a cost benefit analysis of the lockdown in the United Kingdom. International Journal of Clinical Practice 2020.

7. Konig M, Winkler A. COVID-19 and Economic Growth: Does Good Government Performance Pay Off? Inter Econ 2020; 55(4): 224–31.

8. Chen S, Igan D, Pierri N, Presbitero AF. Tracking the Economic Impact of COVID-19 and Mitigation Policies in Europe and the United States. IMF Working Paper 2020; WP/20/125.

9. Zala D, Mosweu I, Critchlow S, Romeo R, McCrone P. Costing the COVID-19 Pandemic: An Exploratory Economic Evaluation of Hypothetical Suppression Policy in the United Kingdom. Value Health 2020; 23(11): 1432–7.

10. Briggs A. Moving beyond ‘lives-saved’ from COVID-19. May 15 2020 https://www.lshtm.ac.uk/research/centres-projects-groups/chil#covid-19 (Accessed 1-July-2020). London School of Hygiene and Tropical Medicine COVID-10 related health economics research 2020.

11. Davies NG, Wikramaratna PS, Clifford S, et al. LSHTM COVID-10 Transmission App https://cmmid.github.io/visualisations/covid-transmission-model (Accessed 17-August-2020). 2020.

12. University of Oxford. CORONAVIRUS GOVERNMENT RESPONSE TRACKER https://www.bsg.ox.ac.uk/research/research-projects/coronavirus-government-response-tracker (Accessed 13-November-2020). 2020.

13. Wikipedia. National responses to COVID-19 pandemic https://en.wikipedia.org/wiki/National_responses_to_the_COVID-19_pandemic (Accessed 19-October-2020). 2020.

14. Davies NG, Kucharski AJ, Eggo RM, Gimma A, Edmunds WJ, Centre for the Mathematical Modelling of Infectious Diseases C-wg. Effects of non-pharmaceutical interventions on COVID-19 cases, deaths, and demand for hospital services in the UK: a modelling study. Lancet Public Health 2020; 5(7): e375–e85.

15. Davies NG, Klepac P, Liu Y, et al. Age-dependent effects in the transmission and control of COVID-19 epidemics. Nat Med 2020; 26(8): 1205–11.

16. Flaxman S, Mishra S, Gandy A, et al. Estimating the effects of non-pharmaceutical interventions on COVID-19 in Europe. Nature 2020; 584(7820): 257–61.

17. Imai N, Cori A, Dorigatti I, et al. Report 3: Transmissibility of 2019-nCoV (https://www.imperial.ac.uk/media/imperial-college/medicine/sph/ide/gida-fellowships/Imperial-College-COVID19-transmissibility-25-01-2020.pdf) (Accessed 1-October-2020). Imperial College London COVID-19 Response Team 25 January 2020 2020.

18. Abbott S, Hellewell J, Thompson R, et al. Estimating the time-varying reproduction number of SARS-CoV-2 using national and subnational case counts [version 1; peer review: awaiting peer review]. Wellcome Open Research 2020; 5(112).

19. PHE. Disparities in the risk and outcomes of COVID-19. https://assets.publishing.service.gov.uk/government/uploads/system/uploads/attachment_data/file/908434/Disparities_in_the_risk_and_outcomes_of_COVID_August_2020_update.pdf (Accessed 29-Sep-2020). Public Health England report 2020.

20. National Institute for Health and Care Excellence. Guide to the methods of technology appraisal. Process and methods guides, http://publicationsniceorguk/pmg9 2013.

21. Worldometer. Countries in the world by population (2020) https://www.worldometers.info/world-population/population-by-country/ (Accessed 3-September-2020). 2020.

22. IMF. World Economic Outlook Update, October 2020: A long and difficult ascent https://www.imf.org/en/Publications/WEO/Issues/2020/09/30/world-economic-outlook-october-2020 (Accessed 7-November-2020). 2020.

23. OECD. Economic Outlook, Volume 2019 Issue 2 https://www.oecd-ilibrary.org/economics/oecd-economic-outlook-volume-2019-issue-2/summary/english_4c90c873-en (Accessed 1-August-2020). 2019.

24. OECD. Purchasing power parities (PPP). https://data.oecd.org/conversion/purchasing-power-parities-ppp.htm (Accessed 5-October-2020). OECD Data 2019.

25. Kulu H, Dorey P. The Contribution of Age Structure to the Number of Deaths from Covid-19 in the UK by Geographical Units. medRxiv 2020.

26. gov.uk. Policy Paper Spending Review 2020. 25th November 2020. https://www.gov.uk/government/publications/spending-review-2020-documents/spending-review-2020 (Accessed 30-Nov-2020). 2020.

27. Baker R, Bloom N, Davis SJ, Terry SJ. COVID-Induced Economic Uncertainty https://www.nber.org/papers/w26983 (Accessed 1-October-2020). National Bureau of Economic Research 2020.

28. Central Statistics Office. National Income and Expenditure 2017: Modified Gross National Income https://www.cso.ie/en/releasesandpublications/ep/p-nie/nie2017/mgni/ (Accessed 19-October-2020). 2017.

29. Leigh S, Granby P. A Tale of Two Thresholds: A Framework for Prioritization within the Cancer Drugs Fund. Value Health 2016; 19(5): 567–76.

## References

1. Mossong J, Hens N, Jit M, et al. Social contacts and mixing patterns relevant to the spread of infectious diseases. PLoS Med 2008; 5(3): e74.

2. Prem K, Cook AR, Jit M. Projecting social contact matrices in 152 countries using contact surveys and demographic data. PLoS Comput Biol 2017; 13(9): e1005697.

3. Briggs A. Moving beyond ‘lives-saved’ from COVID-19. May 15 2020 https://www.lshtm.ac.uk/research/centres-projects-groups/chil#covid-19 (Accessed 1-July-2020). London School of Hygiene and Tropical Medicine COVID-10 related health economics research 2020.

4. PHE. Disparities in the risk and outcomes of COVID-19. https://assets.publishing.service.gov.uk/government/uploads/system/uploads/attachment_data/file/908434/Disparities_in_the_risk_and_outcomes_of_COVID_August_2020_update.pdf (Accessed 29-Sep-2020). Public Health England report 2020.

5. WHO. WHO Coronavirus Disease (COVID-19) Dashboard https://covid19.who.int/region/euro/country/ (Accessed 13-November-2020). 2020.

6. GOV.UK. Coronavirus (COVID-19) in the UK. Cases and deaths in United Kingdom. https://coronavirus-staging.data.gov.uk (Accessed 3-August-2020). 2020.

7. GOV.IE. Updates on COVID-19 (Coronavirus from July-August 2020 https://www.gov.ie/en/publication/b6a9e-updates-on-covid-19-coronavirus-from-july-and-august-2020/ (Accessed 3-August-2020). 2020.

8. Spanish National Centre for Epidemiology. COVID-19 https://cnecovid.isciii.es/covid19/#documentaci%C3%B3n-y-datos (Accessed 3-August-2020). 2020.

9. Tagesspiegel. Interactive data map for sars-cov-2 https://interaktiv.tagesspiegel.de/lab/karte-sars-cov-2-in-deutschland-landkreise/ (Accessed 3-August-2020). 2020.

10. Public Health Agency of Sweden. Covid-19 cases and deaths https://experience.arcgis.com/experience/09f821BQZKqdp2CV3QV5nUEsqSg1ygegLmqRygj (Accessed 3-August-2020). 2020.

11. European Centre for Disease Prevention and Control. Data on hospital and ICU admission rates and current occupancy for COVID-19 https://www.ecdc.europa.eu/en/publications-data/download-data-hospital-and-icu-admission-rates-and-current-occupancy-covid-19 (Accessed 5-October-2020). 2020.

12. ONS. National Life Tables, United Kingdom, 1980-1982 to 2017-2019. https://www.ons.gov.uk/peoplepopulationandcommunity/birthsdeathsandmarriages/lifeexpectancies/datasets/nationallifetablesunitedkingdomreferencetables (Accessed 10-November-2020). 2019.

13. CSO. CSO Statistical release 7 July 2020 11am. Irish Life Tables 2015-2017 https://www.cso.ie/en/releasesandpublications/er/ilt/irishlifetablesno172015-2017/ (Accessed 11-November-2020). 2020.

14. eurostat. Life table https://ec.europa.eu/eurostat/databrowser (Accessed 11-November-2020). 2020.

15. Destatis. Life table (period life table): Germany, period of years, sex, completed age https://www-genesis.destatis.de/genesis/online?sequenz=tabelleErgebnis&selectionname=12621-0001&sachmerkmal=GES&sachschluessel=GESW&language=en#abreadcrumb (Accessed 11-November-2020). 2020.

16. WHO. Global Health Observatory (GHO) data. https://www.who.int/gho/mortality_burden_disease/life_tables/life_tables_text/en/ (Accessed 1-October-2020). 2016.

17. Janssen B, Szende A. Population Norms for the EQ-5D. In: Szende A, Janssen B, Cabases J, eds. Self-Reported Population Health: An International Perspective based on EQ-5D. Dordrecht (NL); 2014: 19–30.

18. Fragaszy EB, Warren-Gash C, White PJ, et al. Effects of seasonal and pandemic influenza on health-related quality of life, work and school absence in England: Results from the Flu Watch cohort study. Influenza Other Respir Viruses 2018; 12(1): 171–82.

19. Hollmann M, Garin O, Galante M, Ferrer M, Dominguez A, Alonso J. Impact of influenza on health-related quality of life among confirmed (H1N1)2009 patients. PLoS One 2013; 8(3): e60477.

20. Docherty AB, Harrison EM, Green CA, et al. Features of 20 133 UK patients in hospital with covid-19 using the ISARIC WHO Clinical Characterisation Protocol: prospective observational cohort study. BMJ 2020; 369: m1985.

21. NHS England. National Cost Collection: National Schedule of NHS costs - Year 2018-19 - NHS trust and NHS foundation trusts. Downloaded from https://www.england.nhs.uk/national-cost-collection/ (Accessed 31-July-2020). 2020.

22. Inensive care national audit & research centre. ICNARC report on COVID-19 in critical care 24 July 2020. 2020.

23. Davies NG, Wikramaratna PS, Clifford S, et al. LSHTM COVID-10 Transmission App https://cmmid.github.io/visualisations/covid-transmission-model (Accessed 17-August-2020). 2020.

24. Omori R, Matsuyama R, Nakata Y. The age distribution of mortality from novel coronavirus disease (COVID-19) suggests no large difference of susceptibility by age. Sci Rep 2020; 10(1): 16642.

25. Folkhalsomyndigheten (Public Health Authority). Antal fall av covid-19 i Sverige – data till och med föregående dag publiceras varje tisdag-fredag https://experience.arcgis.com/experience/09f821BQZKqdp2CV3QV5nUEsqSg1ygegLmqRygj (Accessed 5-August-2020). 2020.

26. Gobierno de Espana. Actualización nº 103. Enfermedad por el coronavirus (COVID-19). 12.05.2020 https://www.mscbs.gob.es/profesionales/saludPublica/ccayes/alertasActual/nCov/documentos/Actualizacion_103_COVID-19.pdf (Accessed 7-November-2020). 2020.

27. statista. Number of coronavirus (COVID-19) deaths in Germany in 2020, by gender and age https://www.statista.com/statistics/1105512/coronavirus-covid-19-deaths-by-gender-germany/ (Accessed 5-August-2020). 2020.

28. University of Oxford. CORONAVIRUS GOVERNMENT RESPONSE TRACKER https://www.bsg.ox.ac.uk/research/research-projects/coronavirus-government-response-tracker (Accessed 13-November-2020). 2020.

29. Our world in data. Total tests performed https://ourworldindata.org/coronavirus-testing#world-map-total-tests-performed-relative-to-the-size-of-population (Accessed 19-October-2020). 2020.

